# Glomerular crescents, IgA-deposits, ANCA, infection – unravelling the diagnostic conundrum

**DOI:** 10.1101/2022.11.13.22281519

**Authors:** Mineaki Kitamura, Salem Almaani, Bindu Challa, Mohankumar Doraiswamy, Isabelle Ayoub, Laura Biederman, Samir V. Parikh, Ana Molovic-Kokovic, Jason Benedict, Nilesh Mhaskar, Zeid Khitan, Sergey V. Brodsky, Tibor Nadasdy, Anjali A Satoskar

## Abstract

**Introduction:** Glomerulonephritis (GN) with crescents and IgA deposits on kidney biopsy poses a frequent diagnostic and therapeutic dilemma because of multiple possibilities.

**Methods:** Native kidney biopsies showing IgA deposition and crescents (excluding lupus nephritis) were identified from our biopsy archives between January 2010 and December 2021. Detailed clinico-pathologic features were assessed. One-year clinical follow-up on a subset of cases was performed.

**Results:** A total of 285 cases were identified and these clustered into IgA nephropathy (IgAN, n=108), Staphylococcus or other infection-associated-GN (SAGN/IRGN, n=46), and anti-neutrophil cytoplasmic antibody associated-GN (ANCA-GN, n=24) based on constellation of clinico-pathologic features, but 101 cases (Group X) could not be definitively differentiated. The reasons have been elucidated, most important being atypical combination of clinico-pathologic features and lack of definitive evidence of active infection. Follow-up (on 72/101 cases), revealed that clinicians’ working diagnosis was IgAN in 42%, SAGN/IRGN in 24%, ANCA-GN in 24%, and others in 7% of the cases, but treatment approach varied from supportive/antibiotics to immunosuppression in each subgroup. Comparing these cases as “received immunosuppression” versus “no-immunosuppression”, only two features - C3-dominant staining; and possibility of recent infection differed (higher in the no-immunosuppression group [p<0.05]). Renal loss was higher in the no-immunosuppression subgroup, but not statistically significant (p=0.11).

**Conclusion:** Diagnostic overlap may remain unresolved in a substantial number of kidney biopsies with glomerular crescents and IgA deposits. A case-by-case approach, appropriate antibiotics if infection is ongoing, and consideration for cautious immunosuppressive treatment for progressive renal dysfunction may be needed for best chance of renal recovery.

## Introduction

Immunoglobulin (Ig) A nephropathy (IgAN) commonly presents with glomerular hematuria and proteinuria. Kidney biopsies of patients with IgAN display IgA-containing immune-complex deposits that are often accompanied with crescent formation. Other glomerulonephritides (GNs) such as Staphylococcus infection-associated GN (SAGN), other causes of infection-related GN (IRGN), IgA vasculitis, and secondary IgAN may also share a similar presentation ^1–8^.

Distinguishing between these entities can be clinically challenging, but is extremely important as the therapeutic approach varies significantly, and ranges from supportive management with SAGN and IRGN to immunosuppression in cases of IgAN ^9,10^. The presence of glomerular crescents adds another layer of complexity as it raises the possibility of antineutrophil cytoplasmic antibodies (ANCA)-associated pauci-immune GN, which despite not being classically associated with immune-complex deposition, can present with mild immunofluorescence staining of IgA and/or IgG, or with a few electron-dense deposits on electron microscopy exam ^11–14^. In addition, around 20% of patient with SAGN/IRGN have positive ANCA serology ^6,15^ and infections (including Staphylococcus aureus infections) can trigger formation of ANCA antibodies (particularly myeloperoxidase (MPO) with resultant vasculitis ^16,17^.

In our previous work, we highlighted some important histologic features distinguishing SAGN from primary IgAN ^6,10,18^. But crescentic cases were not specifically studied. Here, we conducted a retrospective clinico-pathologic analysis on all biopsy cases that showed glomerular IgA deposits accompanied with crescents. The aims of this study were twofold. One was to compare the clinico-pathological features of the three groups of cases that had a definitive etiologic diagnosis on biopsy and build statistical prediction models. The second was to better understand the reasons for the ambiguity in histologic diagnosis of the unclassified cases using clinical follow-up and application of the same prediction models.

## Materials and Methods

### Kidney Biopsy inclusion

Renal biopsy cases showing crescents with glomerular IgA deposits, received at our Medical Center from January 2010 to December 2021 were retrieved from our Renal Pathology files. These included in-house biopsies and biopsies from multiple outside referring hospitals, sent for routine diagnostic work-up. Cases with the diagnosis of lupus nephritis and renal transplant biopsies were excluded.

### Data collection

Demographic data (age, sex, ethnicity); clinical history of infection, laboratory parameters at the time of biopsy (serum creatinine level, serum complement levels (C3 and C4), urine protein, presence/absence of hematuria, blood culture results, and/or other specimen culture results); and serologic data (ANCA testing result) were collected from electronic medical charts and biopsy pathology reports. Our clinical laboratory uses normal cut-offs of >80 mg/dL for C3, >12 mg/dL for C4 ^19^. In some cases, only qualitative results for C3 and C4 (low/normal) were available.

Histopathological features on light microscopy, immunofluorescence staining, and electron microscopy were assessed using the biopsy pathology reports. Exudative glomerular lesions were assessed as previously described ^20,21^. Immunofluorescence staining was assessed semi-quantitatively (trace=0.5+, mild=1+, moderate=2+, strong=3+). The presence or absence of electron-dense immune-type deposits (EDD) on ultrastructural examination was also recorded. When present, the distribution pattern of EDD was evaluated as mesangial only, or mesangial with capillary wall deposits (subendothelial, subepithelial), presence of subepithelial humps, or absence of EDD.

Based on the pathology reports, the cases grouped as follows: Group 1-IgAN, Group 2 - SAGN/IRGN (only included cases with culture-proven infection around the time of GN), Group 3 - ANCA-GN, and Group X - could not be definitively differentiated into any of the three groups based on the biopsy findings and clinical information available to the pathologist at the time of biopsy evaluation. The diagnostic criteria for IgAN, SAGN/IRGN, and ANCA-GN included a combination of clinical, laboratory, and pathologic features as described in reference texts ^6,7,22-26^.

For Group X, a follow-up to understand the management approach used and the status of the renal function at one-year post-biopsy was performed using electronic charts for the cases managed at our Medical Center and by telephone conversations with nephrologists for cases receiving care at outside institutions. Detailed treatment regimens were not available, and renal outcomes were based on the serum creatinine values one year after biopsy and/or the need for dialysis. This study was not meant to be a prospective longitudinal outcomes study, but more directed towards addressing the difficulties in biopsy diagnoses.

### Statistical analysis

Categorical variables were expressed as numbers (percentages), whereas continuous values were expressed as mean±standard deviations or median (interquartile range). Wilcoxon rank sum test, Kruskal-Wallis, and chi-square test were used to assess differences between groups as appropriate. A P-value <0.05 was considered statistically significant. Post-hoc analyses for multiple comparisons were done using the Bonferroni correction. Statistical analyses were performed using the JMP Pro 15.

### Prediction model

Multinomial logistic regression model using a combination of parameters (out of 10 candidate parameters; Supplementary Table 1) was constructed using Groups 1□3 (R version 4.1). Four predictors could be used before the model failed due to excessively wide standard errors. Three best combinations were chosen. A random forest model with 1,000 trees was fit to the data ^27^. To assess the model’s performance, 1000 bootstraps of the data were sampled, the model was refit to each bootstrap, and the mean area under the receiver operating curve (AUROC) was calculated ^28^.

### Ethics

This study was approved by the Ohio State University Internal Review Board (IRB 2011H0364, 2022H0005) and was conducted under the Declaration of Helsinki.

## Results

We had a total of 14,500 kidney biopsies at our facility from January 2010 to December 2021, and 1064 had crescents. Out of these, we identified kidney biopsies showing glomerular crescents and IgA deposition (n=285). The number of patients in Group 1 (IgAN), Group 2 (SAGN/IRGN), Group 3 (ANCA-GN), and Group X (unclassified) were 108, 43, 26, and 101, respectively (Table 1, Supplementary Table 2). There were 7 cases with a diagnosis of IgA vasculitis, but these were not included in our statistical analysis because of the small number.

**Table 1.** Demographic data of patients in all groups and statistical comparisons for Groups 1□3.

|  | **IgAN**<br>**(Group 1)**<br>**(n=108)** | **SAGN/IRGN**<br>**(Group 2)**<br>**(n=43)** | **ANCA-GN**<br>**(Group 3)**<br>**(n=26)** | **P value** | **Unclassified**<br>**(Group X)**<br>**(n=101)** |
| --- | --- | --- | --- | --- | --- |
| **Age (years)<sup>a</sup>** | 40±16 | 53±15* | 59±18* | <0.001 | 60±14 |
| **Age range** | (7 □ 86) | (24 □ 83) | (24 □ 87) |  | (24 □ 91) |
| **Male: Female** | 69:39 | 31:12 | 16:10 | 0.56 | 69:32 |
| **Caucasian** | 80, 83% (n=96) | 34, 94% (n=36) | 17, 81% (n=21) | 0.17 | 73, 86% (n=85) |
| **History of diabetes** | 10, 9% | 13, 30%* | 6, 23% | 0.005 | 27, 27% |
| **History of infection**<br>**(excluding UTI, and**<br>**URTI)** | 0, 0% | 43, 100%*<br>(all culture positive) | 1, 5%† | <0.001 | 29, 29% |
| **ANCA+** | 1, 2% (n=56) | 4, 15% (n=26) | 26, 100%*† (n=26) | <0.001 | 24, 33% (n=73) |
| **sCr (mg/dL) <sup>a</sup>** | 3.1±3.1 | 4.6±2.1* | 5.6±4.1* | <0.001 | 4.7±2.8 |
| **Low C3 (yes)** | 3, 5% (n=57) | 14, 39%* (n=36) | 0, 0%† (n=20) | <0.001 | 12, 14% (n=85) |
| **Low C4 (yes)** | 1, 2% (n=57) | 3, 8% (n=36) | 0, 0% (n=20) | 0.16 | 6, 7% (n=85) |
| **Proteinuria (yes)** | 96, 94% (n=102) | 35, 100% (n=35) | 18, 100% (n=18) | 0.08 | 81, 100% (n=81) |
| **Nephrotic**<br>**proteinuria or >3+** | 49, 48% (n=102) | 21, 60% (n=35) | 3, 17%† (n=18) | 0.008 | 50, 62% (n=81) |
| **Hematuria (yes)** | 86, 91% (n=94) | 34, 94% (n=36) | 22, 96% (n=23) | 0.70 | 79, 96% (n=82) |

2 This is a three-group analysis. (Group X is not included in the statistical analysis.)
<sup>a</sup> Numerical variable. Continuous values are shown with mean±standard deviation or median (interquartile range).
4 Continuous values were analyzed using Kruskal-Wallis test. Categorical values were analyzed using Chi-square test with 5 post-hoc Bonferroni correction (P<0.017 would be significant) \*p<0.017 vs IgAN † p<0.017 vs SAGN/IRGN
6 IgAN, immunoglobulin A nephropathy; SAGN/IRGN, *Staphylococcus* infection–associated glomerulonephritis/ infection 7 related glomerulonephritis; ANCA-GN, antineutrophil cytoplasmic antibodies associated glomerulonephritis; UTI, 8 urinary tract infections; URTI, upper respiratory tract infections

The IgA staining in the ANCA-GN cases was mild and many did not show electron-dense deposits on ultrastructural examination, as highlighted in Table 2, probably representing incidental IgA staining ^26,29,30^. Hence these were diagnosed as ANCA-GN with mild IgA deposits rather than “crescentic IgA nephropathy”. Group 2 (SAGN/IRGN) had culture-proven infection around the time of GN.

**Table 2.**
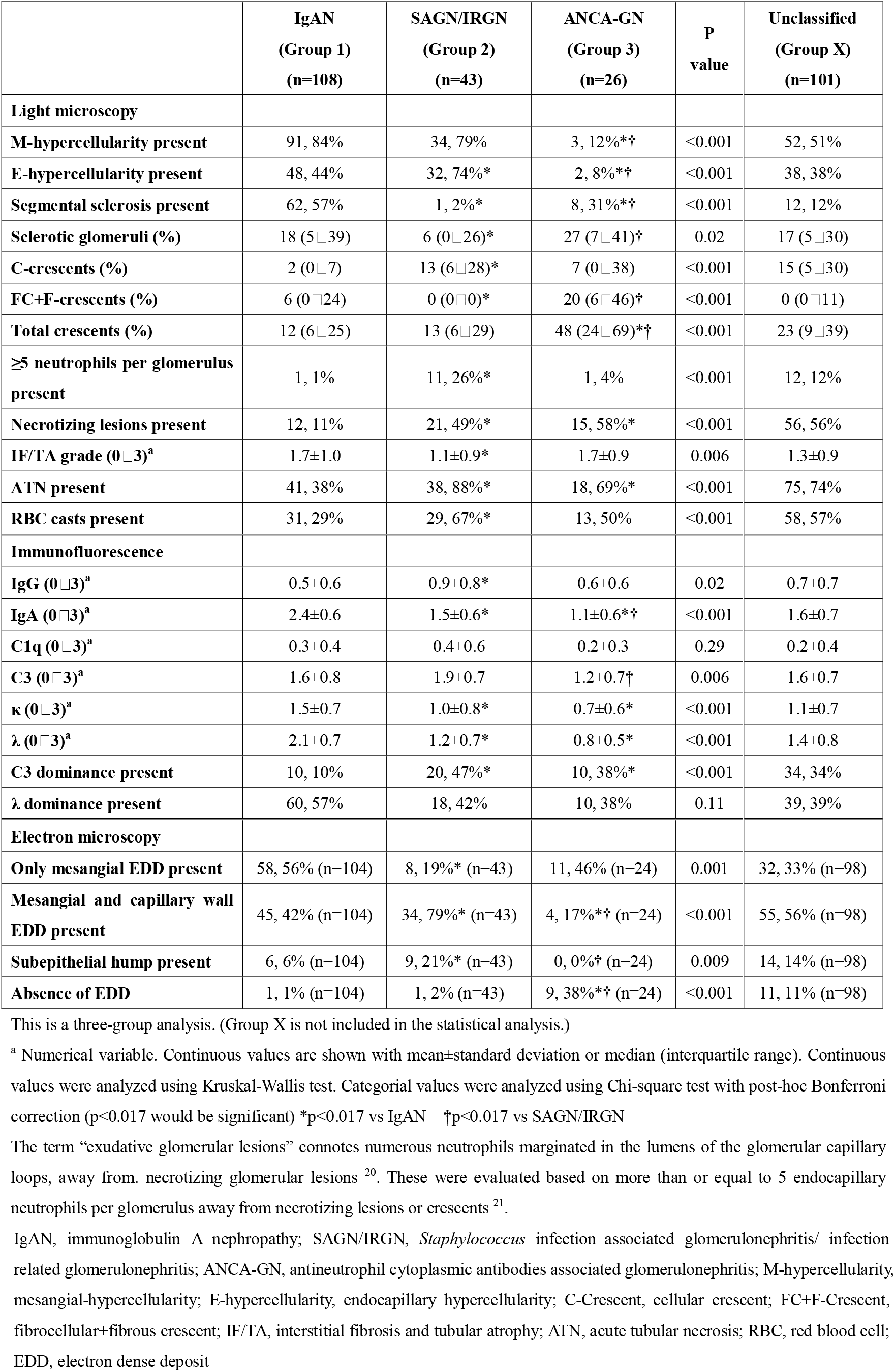
Pathological findings of patients in all groups and statistical comparisons for Groups 1□3.

### Clinico-pathologic features in Groups 1□3

The statistical comparisons are shown in Tables 1 and 2. Group X is shown in the tables but was not included in statistical comparisons with Groups 1□3. Overall, the patients showed a wide age spectrum (7 to 91 years), but mean patient age in SAGN/IRGN and ANCA-GN was significantly higher than in primary IgAN (p<0.001). Significantly higher serum creatinine levels at the time of biopsy were present in SAGN/IRGN and ANCA groups, compared to crescentic IgAN. The SAGN/IRGN group showed the highest percentage of cases with nephrotic range of proteinuria (60%), significantly higher than ANCA-GN (17%) but not compared to IgAN (48%). SAGN did show a significantly higher proportion of patients with low serum C3 compared to those with IgAN (p<0.001). Of 285 cases, 187 underwent ANCA testing, and 55 cases (26 in ANCA-GN, four in SAGN/IRGN, one in IgAN, and 24 in Group X) were ANCA positive. Twenty were proteinase 3 (PR3) positive, 29 were MPO positive, one was dual positive, and five had undefined specificity. The details of infections in cases with SAGN/IRGN are shown in Supplementary Table 3.

Overall, SAGN/IRGN demonstrated features of active glomerular injury (endocapillary hypercellularity, exudative glomerular lesions, higher percent cellular crescents in the biopsy, necrotizing glomerular lesions, acute tubular necrosis [ATN], and tubular red blood cell [RBC] casts), (Fig. 1A, B). In contrast, IgAN showed significantly higher percent fibrocellular/fibrous crescents and the presence of segmental glomerular sclerosis (Fig. 1 C, D) ^5,6,10^. Oxford scores are provided in Supplemental Table 2. ANCA-GN shows a mix of cellular and fibrocellular/fibrous crescents, necrotizing lesions, ATN, and RBC casts as previously described ^11^. Although less frequent, mesangial and endocapillary hypercellularity was noted in 12% and 8% respectively in ANCA-GN. On immunofluorescence (IF), C3 dominance is prevalent in SAGN/IRGN and the reverse in IgAN, as previously reported ^6,10,20^. ANCA-GN showed EDD only in 63% of the biopsies (mainly mesangial) despite presence of mild IgA staining on IF.

**Figure 1:**
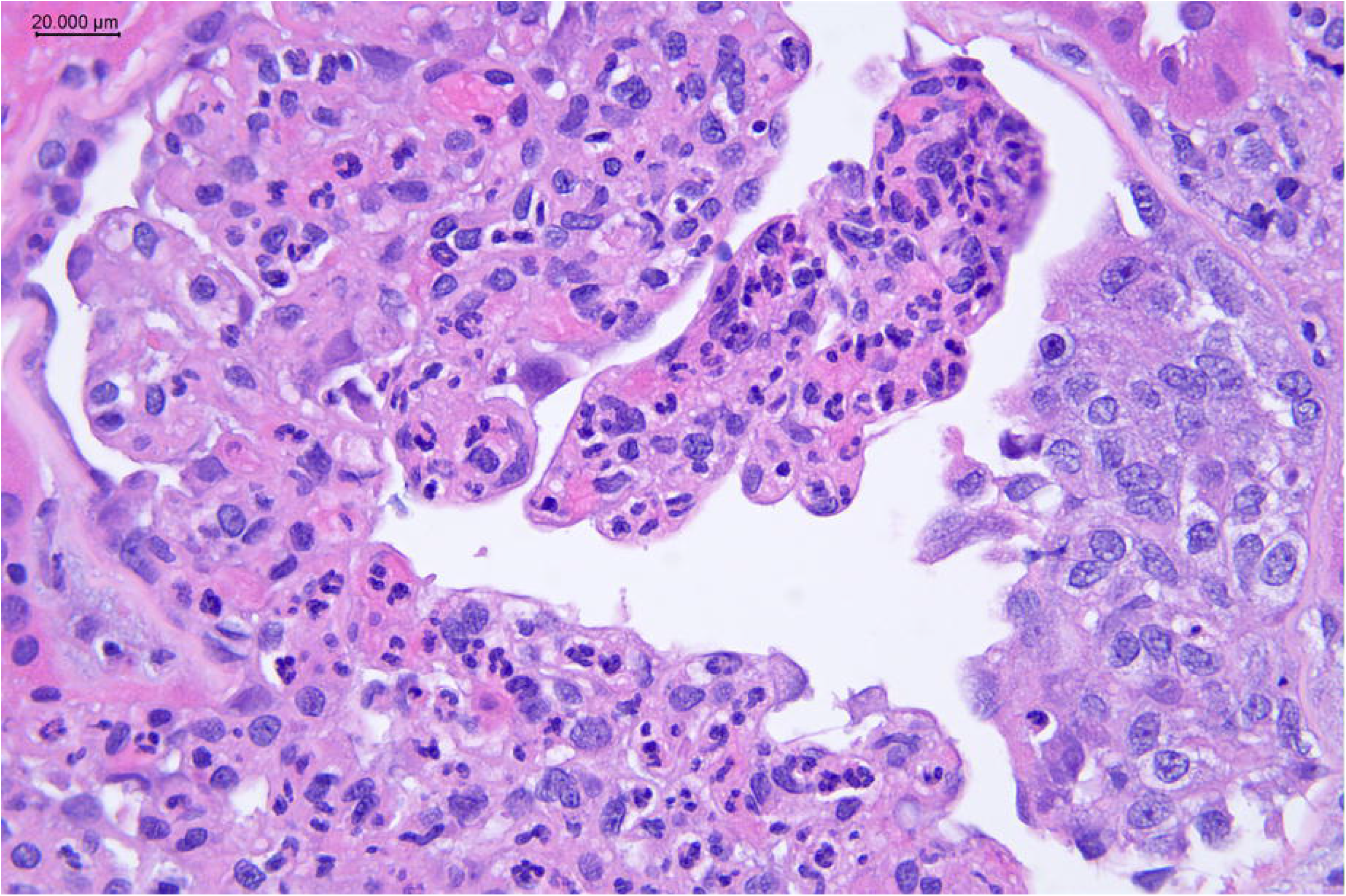
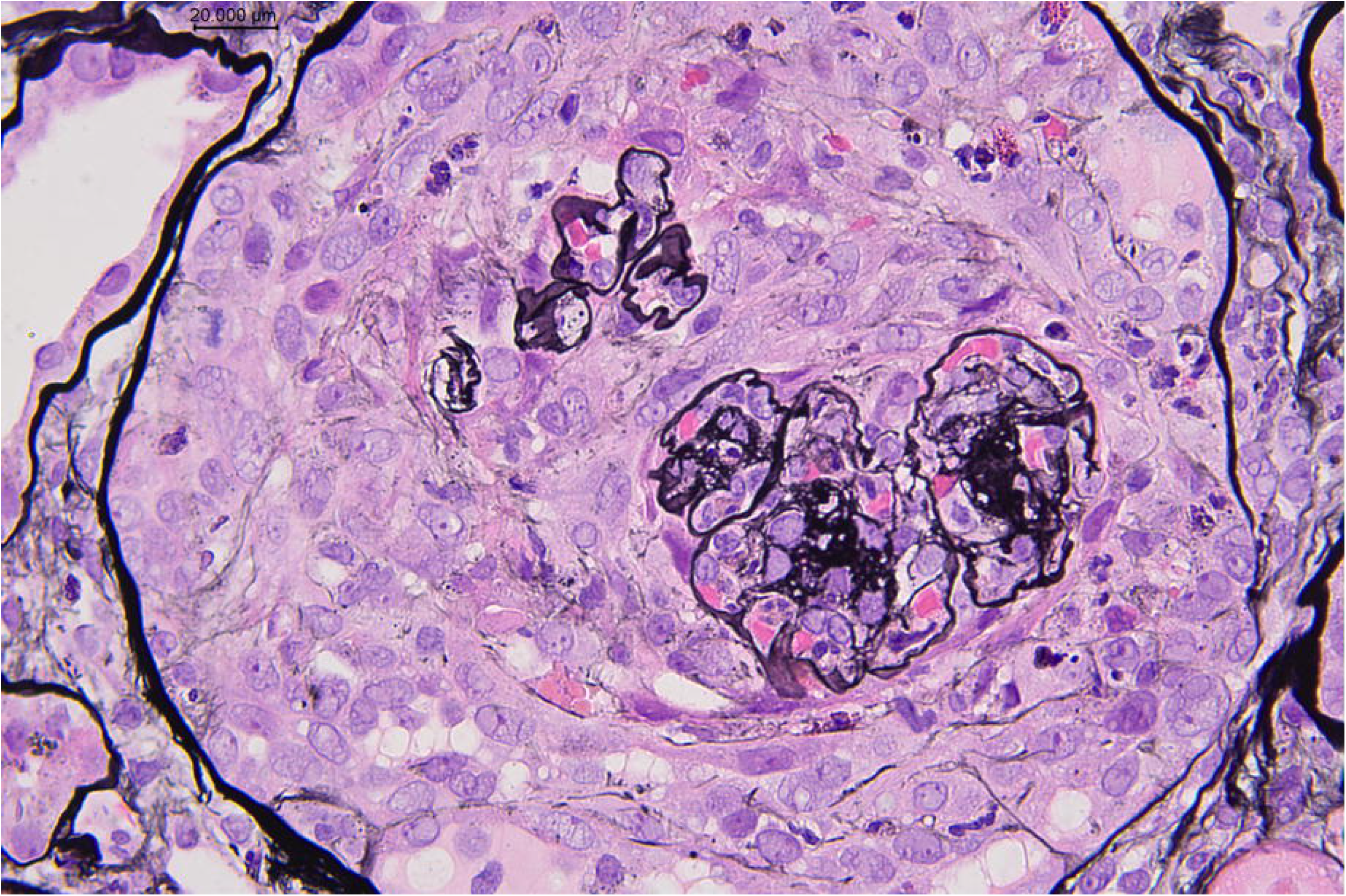
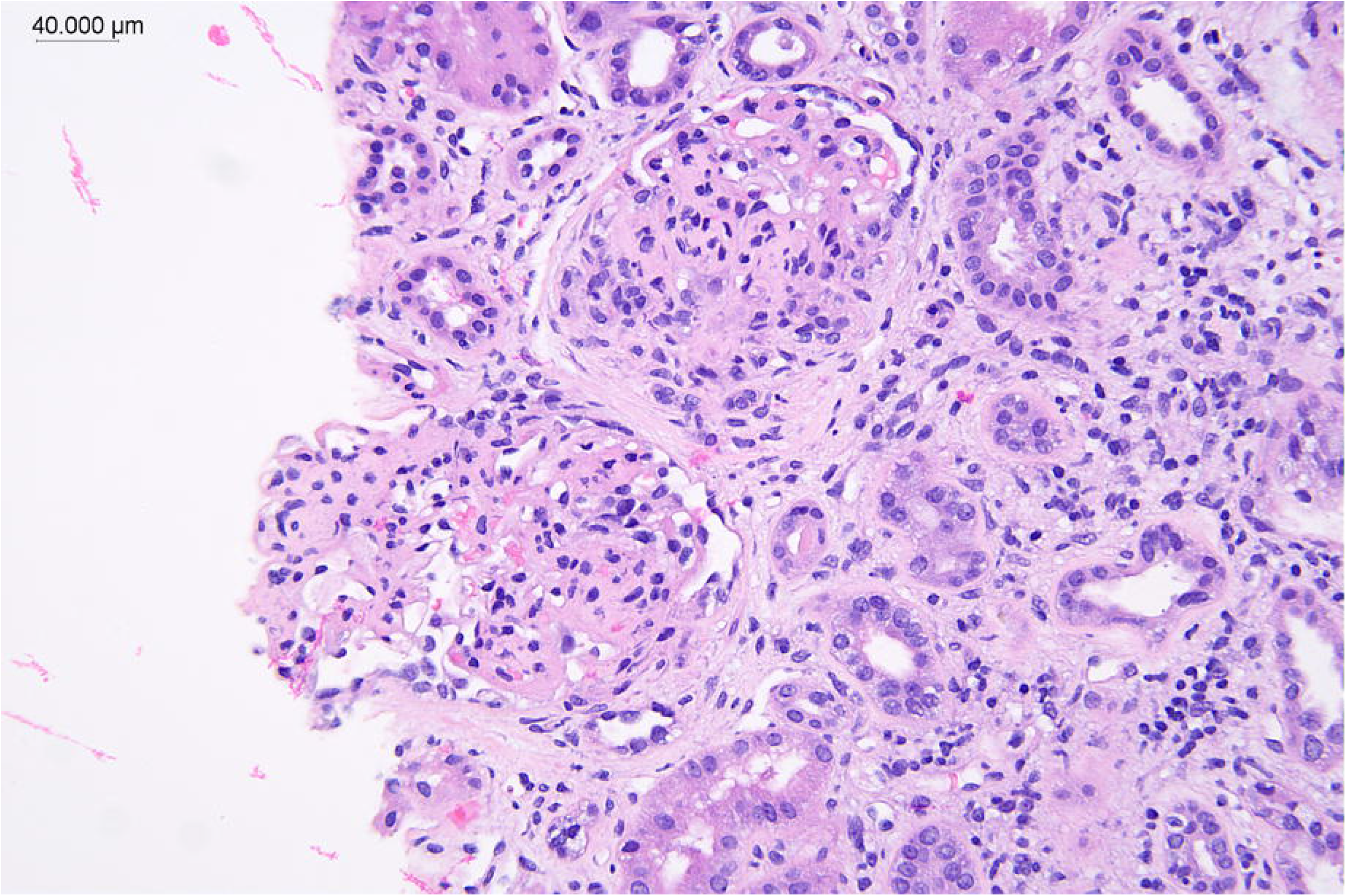
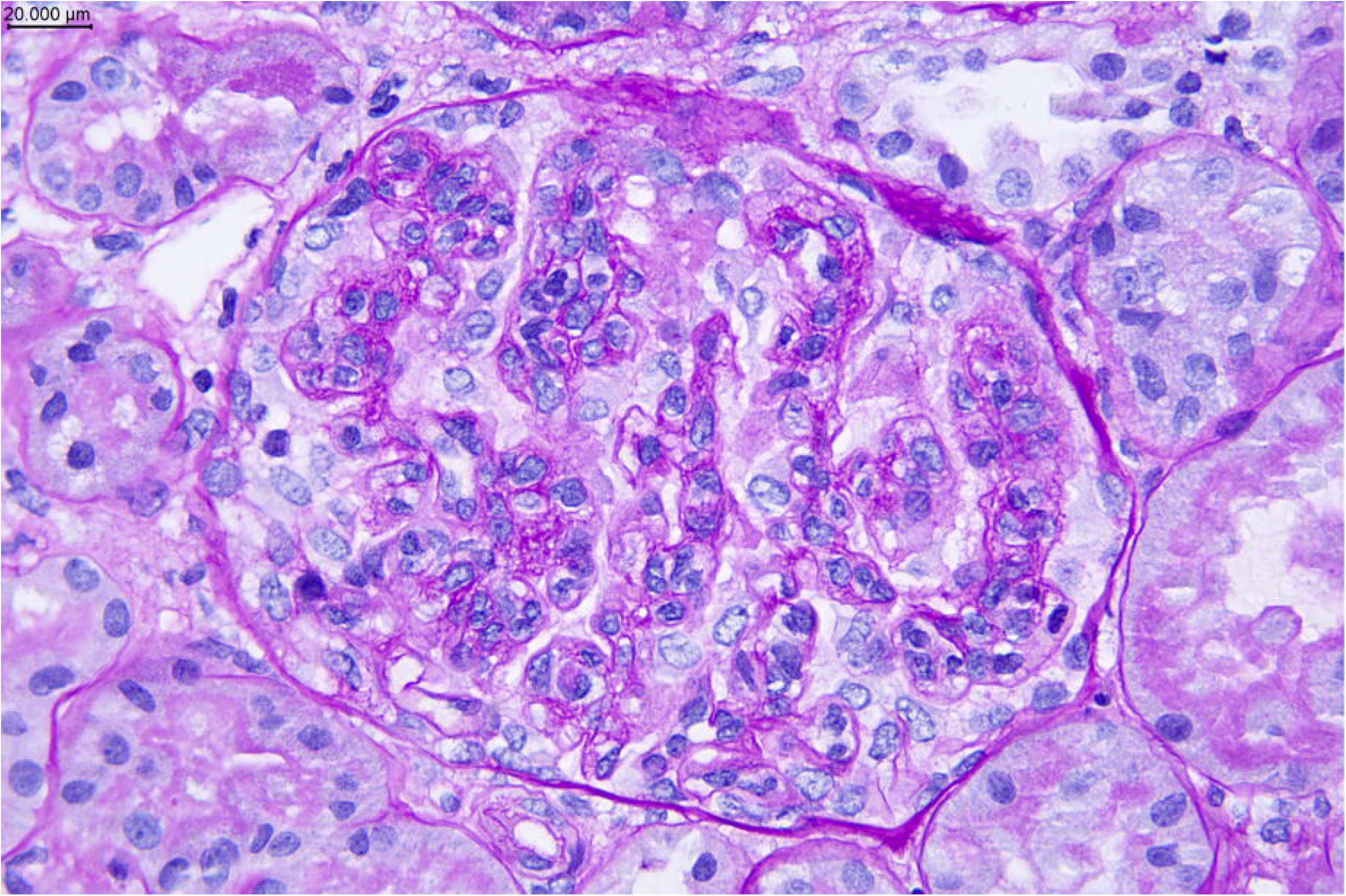
Few characteristic histopathologic features A. Exudative endocapillary hypercellularity in SAGN/IRGN H&E stain 40x; B. Cellular crescent in SAGN/IRGN Jones Methenamine silver stain 40X; C. Segmental sclerosis in IgAN H&E stain 40x; D. Endocapillary hypercellularity in IgAN, Periodic acid Schiff 40x.

### Statistical Prediction models (Groups 1□3)

Three different combinations of parameters (Models 1, 2, 3) gave the best results with the highest AUROC of 0.95 for Groups 1□3 (the accuracy and AUROC values along with the results on bootstraps are shown in Supplementary Tables 4-6, Supplementary Figures 1-3). The confusion matrix and ROC curve for Model 2 are shown in Fig. 2A, B. Model 1 used parameters: ANCA result, IgA intensity, % cellular crescents, segmental sclerosis (present/absent); Model 2 used the same but replaced segmental sclerosis with % fibrocellular crescents; and Model 3 removed ANCA result and replaced with endocapillary hypercellularity (present/absent). Regarding ANCA results, ANCA positive/negative or “not available” were used since ANCA titers were not consistently available.

**Figure 2.**
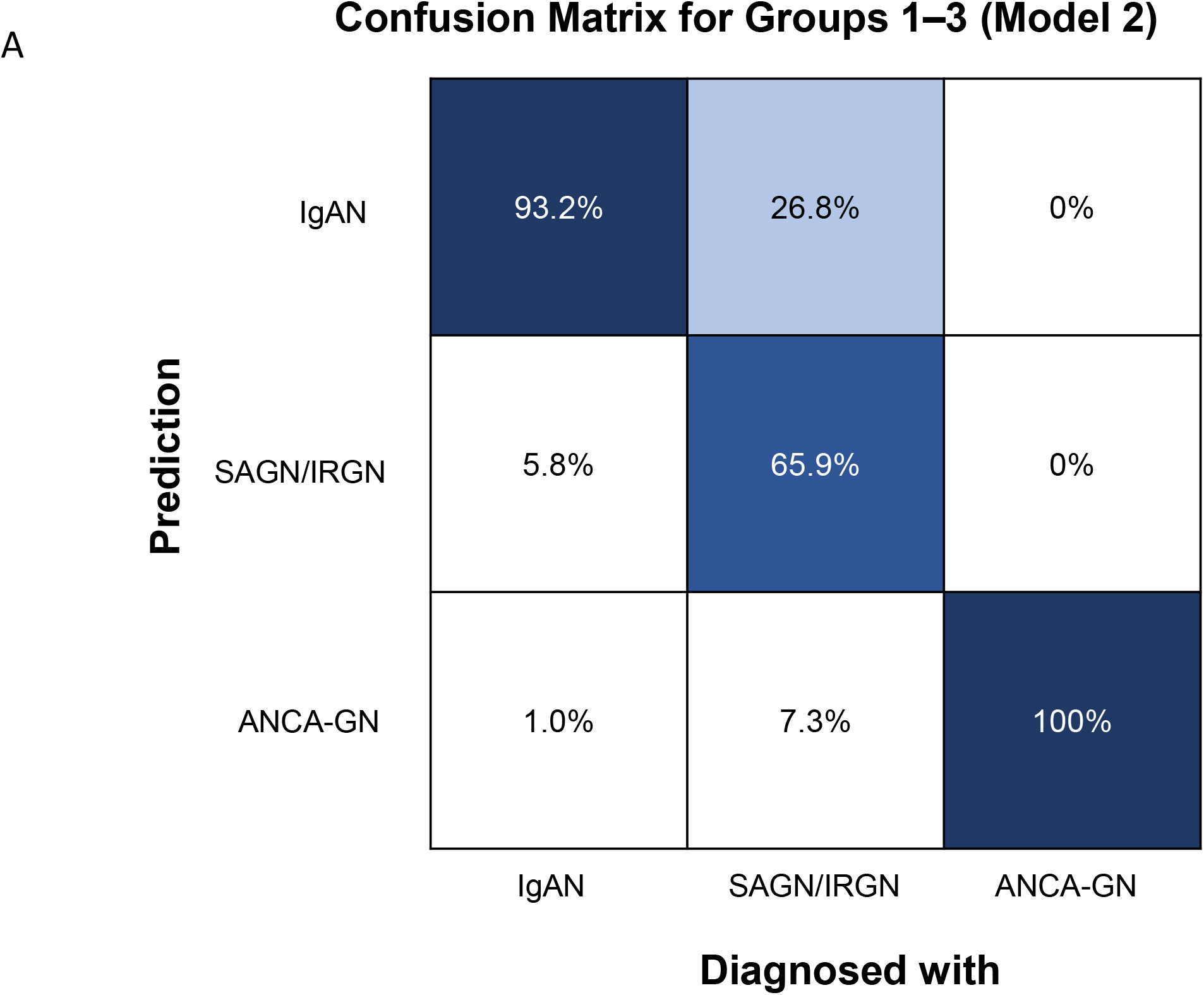
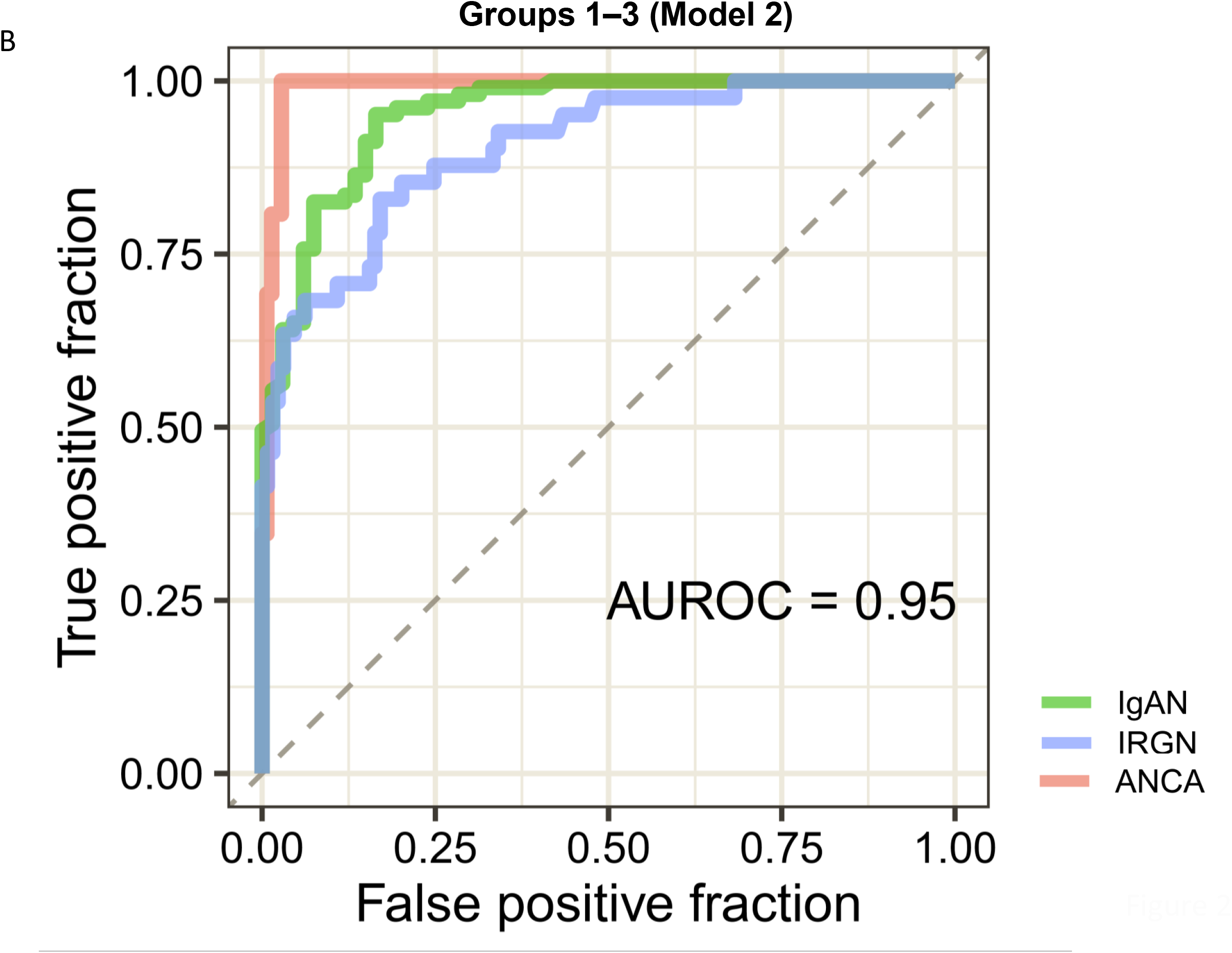
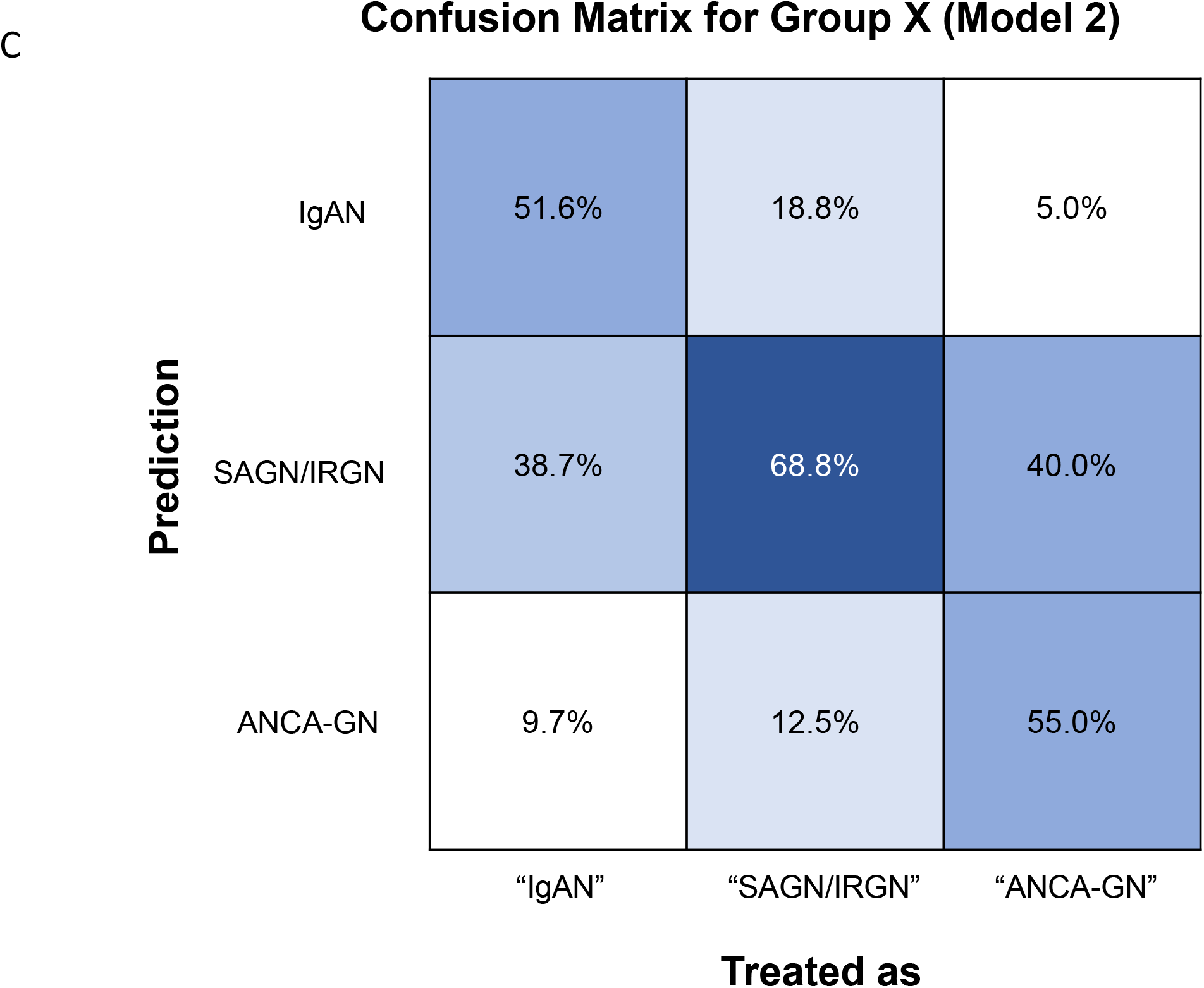
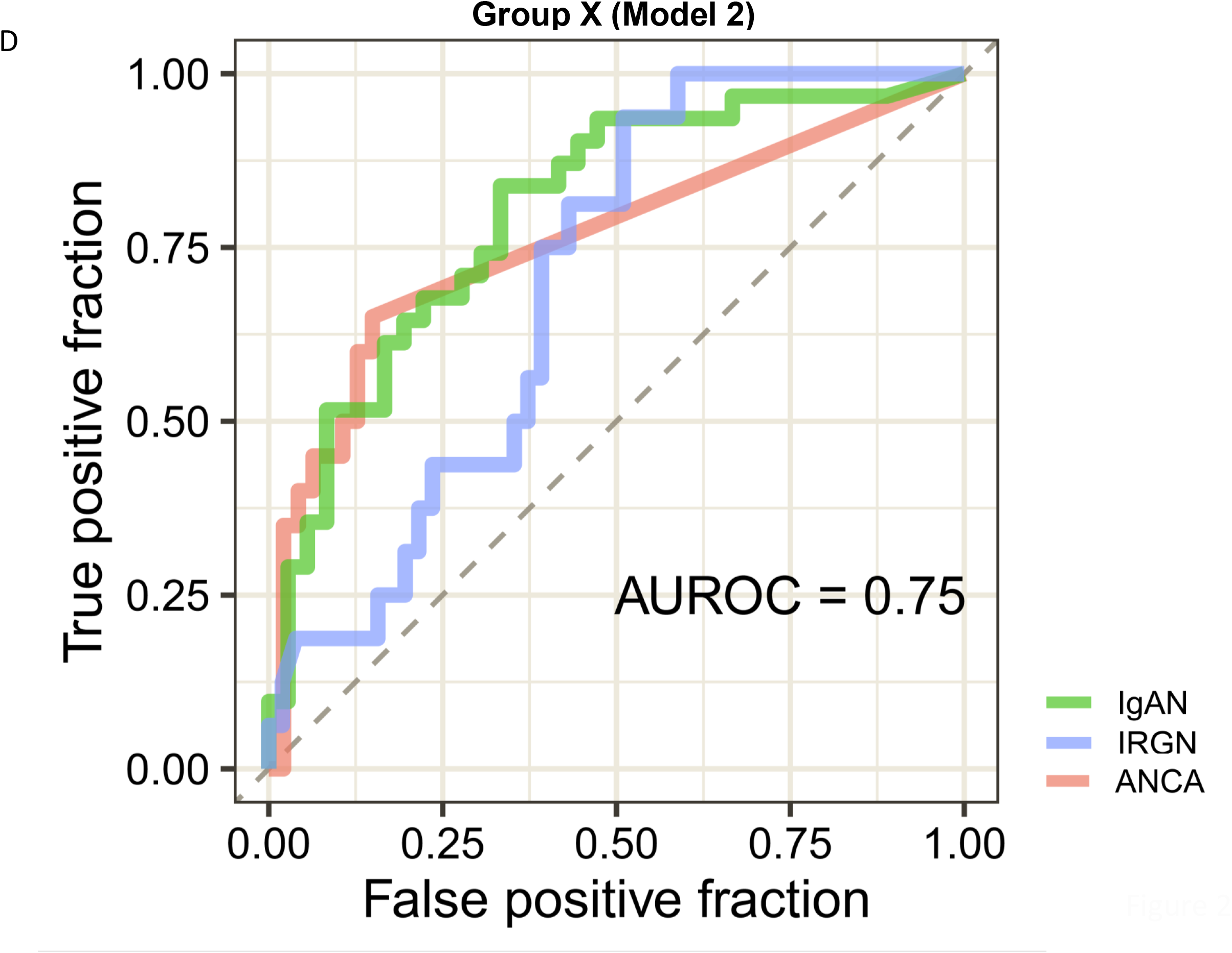
Confusion matrix and receiver operating curve for Model 2 (using parameters ANCA serology, intensity of IgA staining, percent cellular crescents and percent fibrocellular/fibrous crescents. A&B. For Groups 1–3. C&D for Group X.

### Prediction model on Group X patients

Group X constituted 35% of our entire cohort (101/285). Follow-up information was available on 72/101 patients (Table 3). The working diagnosis and management strategy of the treating physician was “IgAN” in 31 (43%), “SAGN/IRGN” in 16 (22%), and “ANCA-GN” in 20 (28%) patients. The remaining 5 (7%) included C3GN (n=1), drug-induced GN (n=1), cryoglobulinemic GN (n=1) and remained unknown (n=2) (Table 3).

**Table 3.** Follow-up and statistical prediction model data on Group X (n=72).

|  | Treated as | “IgAN” | “SAGN/<br>IRGN” | “ANCA-GN” | “Others” | Total |
| --- | --- | --- | --- | --- | --- | --- |
|  | Number (% of total) | 31 (43%) | 16 (22%) | 20 (28%) | 5 (7%) | 72 |
|  | Age (years old) | 59±16 | 60±11 | 60±14 | 62±12 | 60±14 |
|  | Male: Female | 23:8 | 14:2 | 8:12 | 4:1 | 49:23 |
| Patient survival | Living | 23, 74% | 10, 63% | 17, 85% | 4, 80% | 54, 75% |
|  | Deceased | 6, 19% | 5, 31% | 2, 10% | 0, 0% | 13, 18% |
|  | Unknown | 2, 6% | 1, 6% | 1, 5% | 1, 20% | 5, 7% |
| Renal function | Improved or unchanged | 16, 52% | 7, 44% | 12, 60% | 2, 40% | 38, 53% |
|  | Renal loss or worsen | 13, 42% | 7, 44% | 7, 35% | 2, 40% | 28, 39% |
|  | Unknown | 2, 6% | 2, 13% | 1, 5% | 1, 20% | 6, 8% |
| Treatment | Dialysis (including temporary cases) | 15, 48% | 9, 56% | 11, 55% | 2, 40% | 37, 51% |
|  | Antibiotics only | 1, 3% | 10, 63% | 0, 0% | 1, 20% | 12, 17% |
|  | Immunosuppressive therapy | 17, 55% | 0, 0% | 17, 85% | 3, 60% | 37, 51% |
|  | Antibiotics and immunosuppressive therapy | 1, 3% | 4, 25% | 1, 5% | 1, 20% | 6, 8% |
|  | Supportive only | 10, 32% | 2, 13% | 2, 10% | 0, 0% | 14, 19% |
|  | Unknown | 2, 6% | 0, 0% | 0, 0% | 1, 11% | 3, 4% |
| Accuracy of Prediction Models for Group X |  | “IgAN” | “SAGN/<br>IRGN” | “ANCA-GN” | “Others” | AUROC |
| Correctly predicted patients using models | [Model 1]: cellular crescent%, IgA intensity, ANCA serology,<sup>a</sup> and segmental sclerosis | 12, 39% | 9, 56% | 13, 65% | N/A | 0.73 |
|  | [Model 2]: cellular crescent%, IgA intensity, ANCA serology,<sup>a</sup> and fibrous and fibrocellular crescent% | 16, 52% | 11, 69% | 11, 55% | N/A | 0.75 |
|  | [Model 3]: cellular crescent%, IgA intensity, fibrous and fibrocellular crescent%, and endocapillary hypercellularity | 15, 48% | 5, 31% | 14, 70% | N/A | 0.69 |

Others included cryoglobulinemic glomerulonephritis (n=1), C3 glomerulonephritis (n=1), drug-induced glomerulopathy (n=1), and unknown cases (n=2).
Continuous values are shown with mean±standard deviation
IgAN, immunoglobulin A nephropathy; SAGN/IRGN, *Staphylococcus* infection–associated glomerulonephritis/ infection related glomerulonephritis; ANCA, antineutrophil cytoplasmic antibodies associated glomerulonephritis; AUROC, area under the curve of the receiver operating characteristic; N/A, not available
<sup>a</sup> ANCA serology was used at the time of biopsy

The multinomial prediction models were applied to these Group X cases (n=67, after excluding the unclassified 5 cases above). Correlation accuracies of the predicted diagnosis and the working diagnosis of the clinician for all three models are shown in Table 3. Figs. 2 C and D show the results for Model 2. The AUROC was much lower than that seen with Groups 1□3 (and also compared to the bootstrap validation cohorts), emphasizing the difficulty in the biopsy diagnosis of these Group X patients. A summary of the diagnostic pitfalls for Group X is provided in Table 4, based on a re-review of all the pathology reports and the discrepancies in predicted versus clinician’s diagnosis.

**Table 4.** Scenarios precluding definitive biopsy diagnosis and differentiation between ANCA-GN, IgAN and IRGN.

|  |  |
| --- | --- |
| Difficulties in diagnosis of ANCA-GN in Group X | • Crescentic GN with mild IgA staining, ANCA serology pending at the time of biopsy, or negative ANCA serology.<br>• ANCA-GN just preceded by history of disseminated Staphylococcal infection (MRSA or MSSA). Difficult to exclude possibility of remnant foci of infection causing IRGN versus infection-induced ANCA.<br>• Crescentic GN with ANCA positivity, negative culture studies due to difficult to grow bacteria (such as Bartonella). Serology results usually lacking at the time of biopsy.<br>• Presence of mesangial and endocapillary hypercellularity, positive ANCA serology but negative or absent culture results.<br>• Biopsy features resembling ANCA-GN but accompanied by low serum C3 level.<br>• Positive ANCA serology, but strong IgA staining, no evidence of ongoing infection. May be accompanied by leukocytoclastic vasculitic rash. ANCA-GN, crescentic IgAN, IgA vasculitis or occult infection are possibilities.<br>• Past history of autoimmune disease with positive ANA serology or patient on targeted therapies such as tumor necrosis factor blockers and then presents with positive ANCA serology and crescents.<br>• Anti-coagulant related nephropathy (aka Warfarin-related nephropathy) can mimic ANCA-GN with small percentage of crescents in the biopsy and mild IgA deposits. |
| Difficulties in diagnosis of IgAN in Group X | • History of recently treated cellulitis or other infection, negative cultures (but infection reportedly treated before presentation of GN). No prior history of kidney disease, but biopsy shows strong IgA staining with C3.<br>• History of intravenous drug use, but cultures negative, mild proliferative lesions and IgA deposits.<br>• Patient of Asian descent, mild IgA staining with numerous cellular crescents, ANCA serology unavailable at time of biopsy.<br>• Elderly patient with crescents and endocapillary proliferative lesions, nephrotic-range proteinuria, strong C3 staining, but lacking evidence of ongoing infection. |
| Difficulties in diagnosis of SAGN/IRGN in Group X | • Clinical evidence of recent infection (identifiable source) but lack of culture positivity, despite biopsy features suggestive of IRGN.<br>• Recent infection, but past history of kidney biopsy with IgA deposits.<br>• Difficult-to-grow bacteria such as Bartonella, oral flora, cultures negative, serology results lacking.<br>• Cases with weak to absent IgA and C3 staining (pauci-immune pattern), with concomitant crescents and positive ANCA serology.<br>• Mild focal mesangial hypercellularity with crescents and absent endocapillary hypercellularity can mimic ANCA-GN.<br>• Repeated episodes of infection superimposed on underlying co-morbidity such as repeated episodes of pneumonia in patient with chronic obstructive pulmonary disease, or skin infection in patients with bed sores accompanied by persistent proteinuria and hematuria despite antibiotic courses.<br>• Underlying liver cirrhosis, spontaneous bacterial peritonitis, persistent GN with IgA deposits but no positive cultures or definitive clinical site of infection. |

ANCA-GN, antineutrophil cytoplasmic antibodies associated glomerulonephritis; MRSA, methicillin-resistant Staphylococcus aureus; MSSA, methicillin-susceptible Staphylococcus aureus; GN, glomerulonephritis.

Being a retrospective study, treatment approaches in the three main groups in Group X varied and also showed overlap. Therefore, we classified these Group X cases only into 2 subgroups – “those that received immunosuppressive therapy” and “those that received antibiotics/ supportive treatment only”, (Table 5). Cases diagnosed as C3GN, cryoglobulinemic GN, and three cases with unclear follow-up were excluded leaving 67 total patients. On statistical comparison, only C3 dominant immunofluorescence staining, and history of infection (with or without cultures) showed significant differences (p< 0.05). Overall outcomes at one-year post-biopsy are shown in Tables 3 and 4. Renal loss was higher among patients treated without immunosuppression but not statistically significant (p=0.11). Comparing cases with and those without renal failure, only interstitial fibrosis and tubular atrophy showed statistically significant difference (data not shown).

**Table 5.** Clinico-pathologic data of followed-up patients in Group X (n=72) with 1 or without immunosuppression therapy.

|  | Non-immunosuppression<br>(n=26) | Immunosuppression<br>(n=41) | P value |
| --- | --- | --- | --- |
| Age (years)<sup>a</sup> | 61±18 | 59±13 | 0.57 |
| Age range | (31□84) | (29□91) |  |
| Male: Female | 20:6 | 23:18 | 0.08 |
| Caucasian | 19, 90% (n=21) | 31, 89% (n=35) | 0.82 |
| History of diabetes | 7, 27% | 6, 15% | 0.22 |
| History of infection (excluding UTI, and URTI) | 12, 46% | 7, 17% | **0.01** |
| ANCA+ | 5, 31% (n=16) | 17, 47% (n=36) | 0.31 |
| sCr (mg/dL) <sup>a</sup> | 5.2±4.1 | 4.6±2.1 | 0.95 |
| Low C3 (yes) | 5, 23% (n=22) | 3, 9% (n=35) | 0.13 |
| Low C4 (yes) | 1, 5% (n=22) | 2, 6% (n=35) | 0.88 |
| Proteinuria (yes) | 21, 100% (n=21) | 32, 100% (n=32) | 1.00 |
| Nephrotic proteinuria or >3+ | 15, 71% (n=21) | 16, 50% (n=32) | 0.11 |
| Hematuria (yes) | 21, 95% (n=22) | 35, 100% (n=35) | 0.16 |
| Light microscopy |  |  |  |
| M-hypercellularity present | 15, 58% | 20, 49% | 0.48 |
| E-hypercellularity present | 9, 35% | 15, 37% | 0.87 |
| Segmental sclerosis present | 4, 15% | 5, 12% | 0.71 |
| Sclerotic glomeruli (%) | 20 (7□45) | 17 (5□32) | 0.41 |
| C-crescents (%) | 8 (4□23) | 19 (5□36) | 0.18 |
| FC+C-crescents (%) | 0 (0□11) | 0 (0□17) | 0.49 |
| Total crescents (%) | 18 (5□32) | 29 (16□50) | 0.05 |
| ≥5 neutrophils per glomerulus present | 3, 12% | 3, 7% | 0.56 |
| Necrotizing lesions present | 12, 46% | 26, 63% | 0.16 |
| IF/TA grade (0□3)<sup>a</sup> | 1.7±0.7 | 1.5±0.7 | 0.34 |
| ATN present | 17, 65% | 32, 78% | 0.26 |
| RBC casts present | 14, 54% | 27, 66% | 0.33 |
| Immunofluorescence |  |  |  |
| IgG (0□3)<sup>a</sup> | 0.6±0.7 | 0.9±0.8 | 0.11 |
| IgA (0□3)<sup>a</sup> | 1.5±0.6 | 1.7±0.7 | 0.41 |
| C1q (0□3)<sup>a</sup> | 0.3±0.4 | 0.1±0.3 | 0.22 |
| C3 (0□3)<sup>a</sup> | 1.7±0.6 | 1.4±0.8 | 0.02 |
| κ (0□3)<sup>a</sup> | 1.2±0.7 | 1.3±0.8 | 0.56 |
| λ (0□3)<sup>a</sup> | 1.2±0.7 | 1.4±0.8 | 0.17 |
| C3 dominance present | 11, 42% | 8, 20% | **0.045** |
| λ dominance present | 6, 23% | 15, 37% | 0.24 |
| Electron microscopy |  |  |  |
| **Only mesangial EDD present** | 7, 30% (n=23) | 17, 41% (n=41) | 0.38 |
| **Mesangial and capillary wall EDD present** | 14, 61% (n=23) | 20, 49% (n=41) | 0.35 |
| **Subepithelial humps present** | 3, 13% (n=23) | 5, 12% (n=41) | 0.92 |
| **Absence of EDD** | 2, 9% (n=23) | 4, 10% (n=41) | 0.89 |
| **Renal outcomes (improved: worsened)** | 11:14<br>(n=25) | 25:14<br>(n=39) | 0.11 |

C3 glomerulonephritis case (n=1) and cryoglobulinemic glomerulonephritis (n=1) were excluded from this analysis. Renal outcomes were not available in three cases (non-immunosuppression (n=1), immunosuppression (n=2)) <sup>a</sup> Numerical variable. Continuous values are shown with mean±standard deviation or median (interquartile range). Continuous values were analyzed using Wilcoxon sum test. Categorical values were analyzed using Chi-square test (P<0.05 would be significant). UTI, urinary tract infections; URTI, upper respiratory tract infections; ANCA, antineutrophil cytoplasmic antibodies, sCr, serum creatinine level; M-hypercellularity, mesangial-hypercellularity; E-hypercellularity, endothelial hypercellularity; C-Crescent, cellular crescent; FC+F-Crescent, fibrocellular+fibrous crescent; IF/TA, interstitial fibrosis and tubular atrophy; ATN, acute tubular necrosis; RBC, red blood cell; EDD, electron dense deposit

### Trends of infection in Group X patients

As illustrated in the flowchart in Figure 3, Group X contained patients with absent clinical and microbiological evidence of infection; patients that had clinical evidence of infection in the recent past (such as cellulitis, deep-seated abscess, or pneumonia) already treated, but negative/absent culture results; and few cases that did have culture-proven infection but it was already treated with prolonged antibiotic courses, just before presentation of the acute GN. Infection had reportedly resolved just before the onset of acute GN. Subsequent blood cultures were negative and 3/4 of them showed positive ANCA serology.

**Figure 3.**
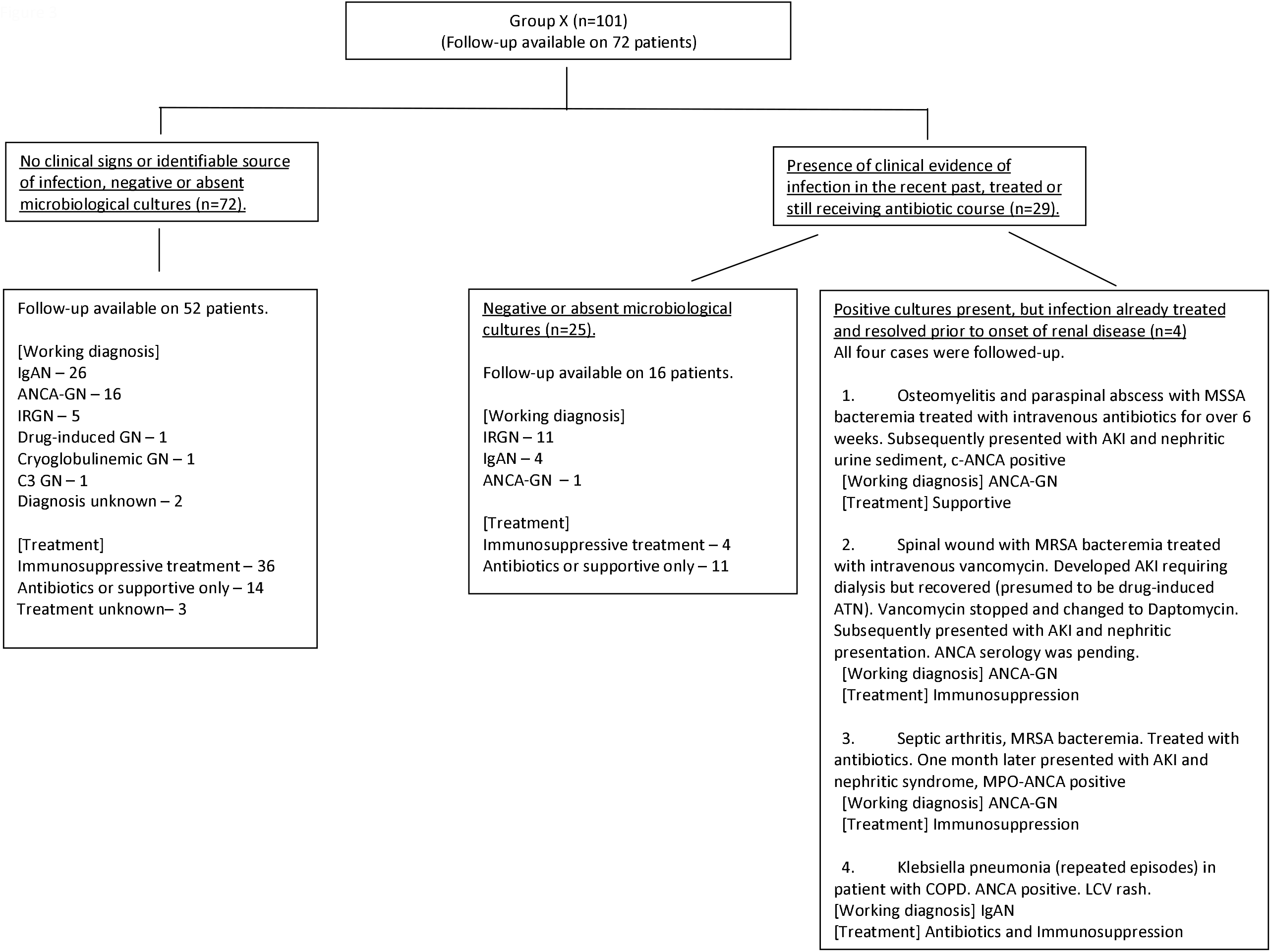
Flow chart showing Group X classified based on history of infection (n=101). Follow-up information about working diagnosis of treating physician, management approach (immunosuppression or not), and kidney function was available for 72/101 patients.

### Clinical management of Group X patients

Among the 16 patients managed as IRGN in Group X (Supplementary Table 7), 4 did develop positive cultures after the biopsy (3 showed MRSA, 1 showed *Bartonella henselae*). Among the 16 presumed SAGN/IRGN patients, most received mainly antibiotics, 4 received antibiotics and steroids (when definite source of infection was not identified). Management for presumed IgAN patients mainly included corticosteroids (up to 6 cases did get cyclophosphamide as well) ^24, 25^. But up to 10 patients were managed only with supportive measures. Management for ANCA-GN included treatment with corticosteroids, and cyclophosphamide (or Rituximab) with or without plasmapheresis ^22, 23^.

## Discussion

This is a retrospective single-center cohort study on patients with biopsies showing glomerulonephritis with crescents and IgA deposits. Differential diagnosis includes crescentic IgAN, SAGN/IRGN and ANCA-GN with incidental IgA deposits. Differentiation between them is critically important. A large number of clinico-pathologic features need to be taken into consideration when evaluating such biopsies to arrive at the correct etiologic diagnosis (Tables 1 and 2) ^6,10,20^. Using these features, majority of the biopsies (184/285 cases, 65%), were accurately differentiated. Each group showed a characteristic constellation of features with good discrimination on the statistical prediction models (AUROC up to 0.95 and 0.93 on bootstrap validation cohorts). However, there was a large subset of biopsies, we called “Group X” (101/285, 35%) in which this differentiation was not possible with certainty because the constellation of clinico-pathologic features was not concordant with either of the three groups (IgAN, SAGN/IRGN or ANCA-GN). The same statistical prediction models when applied and correlated with the clinician’s working diagnosis showed lower accuracy for Group X. Also, a major limitation of these models is that a combination of only a few selected parameters can be used at a time.

Upon dividing the Group X cases based on whether they received immunosuppressive treatment or antibiotics/supportive treatment only (Table 5), none of the clinico-pathologic features showed statistically significant differences between them except for history of recent infection (4 had previous culture positivity, the rest had no microbiological culture results) and C3-dominant immunofluorescence staining (ratio of C3 to IgA intensity greater than 1), highlighting the variability in the management approach among these Group X cases.

Therefore, looking for clinical evidence of infection appears to be most critical in decision-making. Some cases had identifiable source of infection, usually treated or still undergoing treatment, but lacking positive cultures (Fig. 3). Some pathologists may diagnose these as “post-infectious GN in adults” according to criteria described by Nasr et al ^7^. A large percentage of patients in Group X however had neither clinical evidence nor laboratory evidence of infection, despite having biopsy features resembling SAGN/IRGN (high median patient age, high serum creatinine at biopsy, nephrotic-range proteinuria, frequency of active glomerular inflammatory lesions, ATN, and tubular RBC casts). It remains unclear what these cases truly are. These were treated as IgAN by the nephrologist if ANCA serology was negative (although treatment ranged from supportive to administration of immunosuppression), and the remaining were treated as ANCA-GN. Some failed and some recovered (no statistical difference). Few were thought to be IRGN/post-infectious and treated with antibiotics and steroid, but most did not recover. (Fig. 3). The clinico-pathologic features in these cases differ significantly from typical IgAN cases in Group 1 (Supplementary Table 8). Our study thus draws attention to this high-risk subset of presumable IgAN cases (older age, crescentic with nephrotic-range proteinuria) ^31-34^, similar to those described by Sevillano et al. ^35^. They come to attention late in life. Majority of these cases did not have a known history of microscopic hematuria or an established diagnosis of IgAN in the past. These are most likely to be confused with SAGN/IRGN on biopsy. The overall renal outcomes with and without immunosuppression in Group X were not significantly different on retrospective follow-up (Table 5), similar to the findings in the two recent outcomes studies on adult IRGN with and without steroid treatment ^36,37^. But definite conclusions are not possible to draw without large prospective trials. That is difficult to do because most of these patients are elderly with co-morbidities like diabetes mellitus, obesity presenting a high threshold for steroid and immunosuppressive therapy, have varying degree of chronic renal injury on the biopsy, varying extent of glomerular crescents, some have completed antibiotic courses, some are still receiving antibiotics, some patients have stable renal function while some have worsening function, some infectious agents (like *Bartonella*) cannot be detected on culture studies, requiring timely serology. Therefore, treatment decisions have to be taken on an individual basis rather than a predesigned protocol.

The other reasons for diagnostic difficulty revolve around ANCA serology results. Five of 72 followed-up patients in Group X did not have ANCA results at the time of biopsy but were noted to be positive on follow-up. Biopsy sign-out cannot wait until ANCA results are available. In such cases, the decision is up to the nephrologist whether to wait for the ANCA results or start immunosuppressive treatment depending on the extent of activity seen in the biopsy, but occult infection needs to be excluded. Positive ANCA serology in active infection is a known phenomenon. Usually, it is low titer with atypical specificity (lack MPO and PR3 specificity), and often accompanied by mixed serologies such as anti-nuclear antibody, rheumatoid factor. This is particularly common in patients with infective endocarditis ^10,15^. ANCA positivity in patients with strong IgA staining and EDD in the absence of clinical and laboratory evidence of infection, can pose a diagnostic dilemma (“crescentic IgAN with incidental ANCA positivity” versus “ANCA-GN with IgA deposits”). In such cases, rather than struggling for the “correct diagnosis”, focusing on immunosuppressive treatment is more crucial and appropriate.

Another important cause for ambiguity in biopsy diagnosis is when there is culture-proven disseminated Staphylococcal infection just before the onset of ANCA-positive GN (Fig. 3). The infection was reportedly treated with prolonged antibiotic courses before onset of GN. This raises the vexing question of whether this is truly IRGN/post-infectious GN with positive ANCA serology or is it ANCA-GN triggered by Staphylococcal antigens released during the infection, due to uncontrolled neutrophilic activation, ineffective apoptosis and neutrophil extracellular traps, as described for idiopathic ANCA-GN ^16,17^. At what point can ANCA-positive infection-associated GN be safely excluded and a diagnosis of ANCA-GN be rendered can be a difficult issue ^38^. One such patient (Patient 1 in Fig. 3) refused steroids and cyclophosphamide-based treatment regimen, and soon progressed to end-stage renal disease. Another patient (Patient 3), on the other hand, was treated with immunosuppression regimen and did recover renal function. Therefore, carefully considering the temporal relation between onset of GN in relation to the infection and treating accordingly is very important. Another difficult situation arises in patients with repeated episodes of infection superimposed on underlying co-morbidity (such as pneumonia in chronic lung disease in Patient 4 Fig. 3). Despite repeated courses of antibiotics, there may be persistent nephritic kidney disease. At some point, steroid treatment and even cyclophosphamide may be needed, tailored to the underlying condition of the patient ^39, 40^. Several other difficult diagnostic scenarios found in Group X are summarized in Table 4. In cases with dual MPO and PR3 positive serology, the possibility of drug-induced GN is a consideration ^41,42^.

Five patients in this entire cohort of 285 patients had advanced liver cirrhosis. Such cases provide another diagnostic challenge. Microbiological cultures may be negative. The cause of the GN in such cases often remains unclear. It could be multifactorial with severe disturbance in gut-liver axis and lack of clearance of immune-complex deposits from blood ^3,4,43,44^. Also, these patients tend to be critically ill and tend to have a poor prognosis irrespective of immunosuppressive treatment or antibiotic therapy alone.

This is a retrospective single-center study and has few pitfalls. Retrieving clinical follow-up for biopsies received from outside hospitals is challenging. Treatment approaches varied between treating physicians and patient decisions, therefore correlation of pathologic diagnoses with outcomes was not possible with this study design. Also, clinical outcomes are influenced by patient age, associated co-morbidities, and underlying chronic renal injury. ANCA titers were not consistently available. Only ANCA positive/negative results were used. Complement levels were missing in a sizeable portion of the patients (particularly in cases with IgAN). For our multinomial regression prediction models, we did perform internal validation using 1000 bootstraps from our dataset itself, however, it lacks a separate validation cohort. Patients in this study were mainly Caucasian, but geographical differences in prevalence and severity of IgAN do exist ^1,2,11^.

In summary, definitive etiologic diagnosis on biopsies with crescentic GN and concomitant IgA deposits may not be possible in a subset of cases even after evaluating all the histologic features described. The most important thing is to exclude ongoing/occult infection (based on clinical signs, imaging studies, culture studies and also serologic studies for fastidious bacteria, harboring a niche for infection such as prosthetic devices, artificial heart valves, intravenous drug use). Source of infection however may not be present in every case. If acute ongoing infection (or even chronic infection as in diabetic patients) can be safely excluded and/or has been adequately treated and biopsy shows extensive active crescents and worsening renal function, use of immunosuppressive treatment tailored to the underlying condition of the patient may be beneficial, as they are at high risk for disease progression. If crescents are few and segmental and renal dysfunction is not progressing (as in “post-infectious” GN), then a brief period of observation to see if it resolves, may also be reasonable. Standardized treatment protocols are difficult to design. Decision needs to be made on a case-by-case basis in these elderly patients with co-morbidities. Treatment for IRGN and post-infectious GN in the elderly remains challenging ^36,37^. Outcomes are difficult to predict. Targeted therapies against dysregulated neutrophilic activation may be an option to explore ^45^.

## Data Availability

All data produced in the present work are contained in the manuscript and supplemental data.

## Acknowledgments

We acknowledge all our referring Nephrologists for the detailed clinical follow-up information on their patients, specifically mentioning Dr. Darren Dreyfus, Dr. Sreedevi Chennupati, Dr. Kevin Pargeter, Dr. Sunil Akkina, Dr. Ahsan Baig, Dr. Joshua Bitter, and Dr. Christopher Brown.

## Sources of Funding

None

## Disclosures

None

## Notes

### Competing Interest Statement

The authors have declared no competing interest.

### Funding Statement

This study did not receive any funding

### Author Declarations

The Ohio State University Wexner Medical Center IRB 2011H0364 IRB 2022H0005

## References

1. Barratt J, Feehally J. IgA nephropathy. J Am Soc Nephrol. 2005;16(7):2088–2097. doi:10.1681/ASN.2005020134

2. Jennette JC, Olson JL, Silva FG, D’Agati VD. Heptinstall’s pathology of the kidney. 7 th ed. 2015 Chapter 12.

3. Saha MK, Julian BA, Novak J, Rizk D V. Secondary IgA nephropathy. Kidney Int. 2018;94(1):674–681. doi:10.1016/j.kint.2018.02.030

4. Hemminger J, Arole V, Ayoub I, Brodsky S V., Nadasdy T, Satoskar AA. Acute glomerulonephritis with large confluent IgA-dominant deposits associated with liver cirrhosis. PLoS One. 2018;13(4):e0193274. doi:10.1371/journal.pone.0193274

5. Satoskar AA, Nadasdy G, Plaza JA, et al. Staphylococcus infection-associated glomerulonephritis mimicking IgA nephropathy. Clin J Am Soc Nephrol. 2006;1(6):1179–1186. doi:10.2215/CJN.01030306

6. Satoskar AA, Suleiman S, Ayoub I, et al. Staphylococcus infection–associated GN – spectrum of IgA staining and prevalence of ANCA in a single-center cohort. Clin J Am Soc Nephrol. 2017;12(1):39–49. doi:10.2215/CJN.05070516

7. Nasr SH, Radhakrishnan J, D’Agati VD. Bacterial infection-related glomerulonephritis in adults. Kidney Int. 2013;83(5):792–803. doi:10.1038/ki.2012.407

8. Alexander S, Yusuf S, Rajan G, et al. Crescentic glomerulonephritis: What’s different in South Asia? A single center observational cohort study. Wellcome Open Res. 2020;5:164. doi:10.12688/wellcomeopenres.16071.1

9. Satoskar AA, Molenda M, Shim R, et al. Henoch-Schönlein purpura-like presentation in IgA-dominant staphylococcus infectionassociated glomerulonephritis - A diagnostic pitfall. Clin Nephrol. 2013;79(4):302–312. doi:10.5414/CN107756

10. Satoskar AA, Parikh S V., Nadasdy T. Epidemiology, pathogenesis, treatment and outcomes of infection-associated glomerulonephritis. Nat Rev Nephrol. 2020;16(1):32–50. doi:10.1038/s41581-019-0178-8

11. Jennette JC. Rapidly progressive crescentic glomerulonephritis. Kidney Int. 2003;63(3):1164–1177. doi:10.1046/j.1523-1755.2003.00843.x

12. Haas M, Jafri J, Bartosh SM, Karp SL, Adler SG, Meehan SM. ANCA-associated crescentic glomerulonephritis with mesangial IgA deposits. Am J Kidney Dis. 2000;36(4):709–718. doi:10.1053/ajkd.2000.17615

13. Haas M, Eustace JA. Immune complex deposits in ANCA-associated crescentic glomerulonephritis: A study of 126 cases. Kidney Int. 2004;65(6):2145–2152. doi:10.1111/j.1523-1755.2004.00632.x

14. Neumann I, Regele H, Kain R, Birck R, Meisl FT. Glomerular immune deposits are associated with increased proteinuria in patients with ANCA-associated crescentic nephritis. Nephrol Dial Transplant. 2003;18(3):524–531. doi:10.1093/NDT/18.3.524

15. Boils CL, Nasr SH, Walker PD, Couser WG, Larsen CP. Update on endocarditis-associated glomerulonephritis. Kidney Int. 2015;87(6):1241–1249. doi:10.1038/ki.2014.424

16. Kakoullis L, Parperis K, Papachristodoulou E, Panos G. Infection-induced myeloperoxidase specific antineutrophil cytoplasmic antibody (MPO-ANCA) associated vasculitis: A systematic review. Clin Immunol. 2020;220(August):108595. doi:10.1016/j.clim.2020.108595

17. Miranda-Filloy JA, Veiga JA, Juarez Y, Gonzalez-Juanatey C, Gonzalez-Gay MA, Garcia-Porrua C. Microscopic polyangiitis following recurrent Staphylococcus aureus bacteremia and infectious endocarditis. Clin Exp Rheumatol. 2006;24(6):705–706.

18. Brodsky S V., Nadasdy T, Cassol C, Satoskar A. IgA Staining Patterns Differentiate Between IgA Nephropathy and IgA-Dominant Infection-Associated Glomerulonephritis. Kidney Int Reports. 2020;5(6):909–911. doi:10.1016/j.ekir.2020.03.029

19. Birmingham DJ, Irshaid F, Nagaraja HN, et al. The complex nature of serum C3 and C4 as biomarkers of lupus renal flare. Lupus. 2010;19(11):1272–1280. doi:10.1177/0961203310371154

20. Jennette JC, Olson JL, Silva FG, D’Agati VD. Heptinstall’s pathology of the kidney. 7 th ed. 2015 Chapter 10.

21. Haas M, Racusen LC, Bagnasco SM. IgA-dominant postinfectious glomerulonephritis: a report of 13 cases with common ultrastructural features. Hum Pathol. 2008;39(9):1309–1316. doi:10.1016/j.humpath.2008.02.015

22. Molnár A, Studinger P, Ledó N. Diagnostic and Therapeutic Approach in ANCA-Associated Glomerulonephritis: A Review on Management Strategies. Front Med. 2022;9(June): 884188. doi:10.3389/fmed.2022.884188

23. Jain K, Jawa P, Derebail VK, Falk RJ. Treatment Updates in Antineutrophil Cytoplasmic Autoantibodies (ANCA) Vasculitis. Kidney360. 2021;2(4):763–770. doi:10.34067/kid.0007142020

24. Roccatello D, Ferro M, Cesano G, et al. Steroid and cyclophosphamide in IgA nephropathy. Nephrol Dial Transplant. 2000;15(6):833–835. doi:10.1093/ndt/15.6.833

25. Gutiérrez E, Carvaca-Fontán F, Luzardo L, Morales E, Alonso M, Praga M. A Personalized Update on IgA Nephropathy: A New Vision and New Future Challenges. Nephron. 2020;144(11):555–571. doi:10.1159/000509997

26. Berden AE, Ferrario F, Hagen EC, et al. Histopathologic classification of ANCA-associated glomerulonephritis. J Am Soc Nephrol. 2010;21(10):1628–1636. doi:10.1681/ASN.2010050477

27. Greenwell BM, Boehmke BC. Variable Importance Plots—An Introduction to the vip Package. R J. 2020;12(1):343–366. doi:10.32614/rj-2020-013

28. Hand DJ, Till RJ. A Simple Generalisation of the Area Under the ROC Curve for Multiple Class Classification Problems. Mach Learn. 2001;45(2):171–186. doi:10.1023/A:1010920819831

29. Sinniah R. Occurrence of mesangial IgA and IgM deposits in a control necropsy population. J Clin Pathol. 1983;36(3):276–279. doi:10.1136/jcp.36.3.276

30. Suzuki K, Honda K, Tanabe K, Toma H, Nihei H, Yamaguchi Y. Incidence of latent mesangial IgA deposition in renal allograft donors in Japan. Kidney Int. 2003;63(6):2286–2294. doi:10.1046/j.1523-1755.63.6s.2.x

31. Haas M, Verhave JC, Liu ZH, et al. A multicenter study of the predictive value of crescents in IgA nephropathy. J Am Soc Nephrol. 2017;28(2):691–701. doi:10.1681/ASN.2016040433

32. Trimarchi H, Barratt J, Cattran DC, et al. Oxford Classification of IgA nephropathy 2016: an update from the IgA Nephropathy Classification Working Group. Kidney Int. 2017;91(5):1014–1021. doi:10.1016/j.kint.2017.02.003

33. Chen CH, Wu MJ, Wen MC, Tsai SF. Crescents formations are independently associated with higher mortality in biopsy-confirmed immunoglobulin A nephropathy. PLoS One. 2020;15(7 July): e0237075. doi:10.1371/journal.pone.023707

34. Tanaka S, Ninomiya T, Katafuchi R, et al. Development and validation of a prediction rule using the Oxford classification in IgA nephropathy. Clin J Am Soc Nephrol. 2013;8(12):2082–2090. doi:10.2215/CJN.03480413

35. Sevillano AM, Diaz M, Caravaca-Fontán F, et al. IgA nephropathy in elderly patients. Clin J Am Soc Nephrol. 2019;14(8):1183–1192. doi:10.2215/CJN.13251118

36. Arivazhagan S, Lamech TM, Myvizhiselvi M, et al. Efficacy of Corticosteroids in Infection-Related Glomerulonephritis-A Randomized Controlled Trial. Kidney Int reports. 2022;7(10):2160–2165. doi:10.1016/J.EKIR.2022.07.163

37. John EE, Thomas A, Eapen JJ, et al. Latency, Anti-Bacterial Resistance Pattern, and Bacterial Infection-Related Glomerulonephritis. Clin J Am Soc Nephrol. 2021; 16: 1210–1220. 2021;16(8):1210-1220. doi:10.2215/CJN.18631120

38. Mohandes S, Satoskar A, Hebert L, Ayoub I. Bacterial endocarditis manifesting as autoimmune pulmonary renal syndrome: ANCA-associated lung hemorrhage and pauci-immune crescentic glomerulonephritis. Clin Nephrol. 2018;90(6):431–433. doi:10.5414/CN109495

39. Nasr SH, Markowitz GS, Stokes MB, Said SM, Valeri AM, D’Agati VD. Acute postinfectious glomerulonephritis in the modern era: Experience with 86 adults and review of the literature. Medicine (Baltimore). 2008;87(1):21–32. doi:10.1097/md.0b013e318161b0fc

40. Roy S, Murphy WM, Arant BS. Poststreptococcal crescenteric glomerulonephritis in children: Comparison of quintuple therapy versus supportive care. J Pediatr. 1981;98(3):403–410. doi:10.1016/S0022-3476(81)80703-2

41. Santoriello D, Bomback AS, Kudose S, et al. Anti-neutrophil cytoplasmic antibody associated glomerulonephritis complicating treatment with hydralazine. Kidney Int. 2021;100(2):440–446. doi:10.1016/j.kint.2021.03.029

42. Hogan JJ, Markowitz GS, Radhakrishnan J. Drug-induced glomerular disease: Immune-mediated injury. Clin J Am Soc Nephrol. 2015;10(7):1300–1310. doi:10.2215/CJN.01910215

43. Raj D, Tomar B, Lahiri A, Mulay SR. The gut-liver-kidney axis: Novel regulator of fatty liver associated chronic kidney disease. Pharmacol Res. 2020;152(December 2019):104617. doi:10.1016/j.phrs.2019.104617

44. Lehto M, Groop PH. The gut-kidney axis: Putative interconnections between gastrointestinal and renal disorders. Front Endocrinol (Lausanne). 2018;9(SEP): 553. doi:10.3389/fendo.2018.00553

45. Terui H, Yamasaki K, Wada-Irimada M, et al. Staphylococcus aureus skin colonization promotes SLE-like autoimmune inflammation via neutrophil activation and the IL-23/IL-17 axis. Science immunology 2022; 7: eabm9811. doi:10.1126/sciimmunol.abm9811

