## Supplementary figures for "Glomerular crescents, IgA-deposits, ANCA, infection – unravelling the diagnostic conundrum"

A

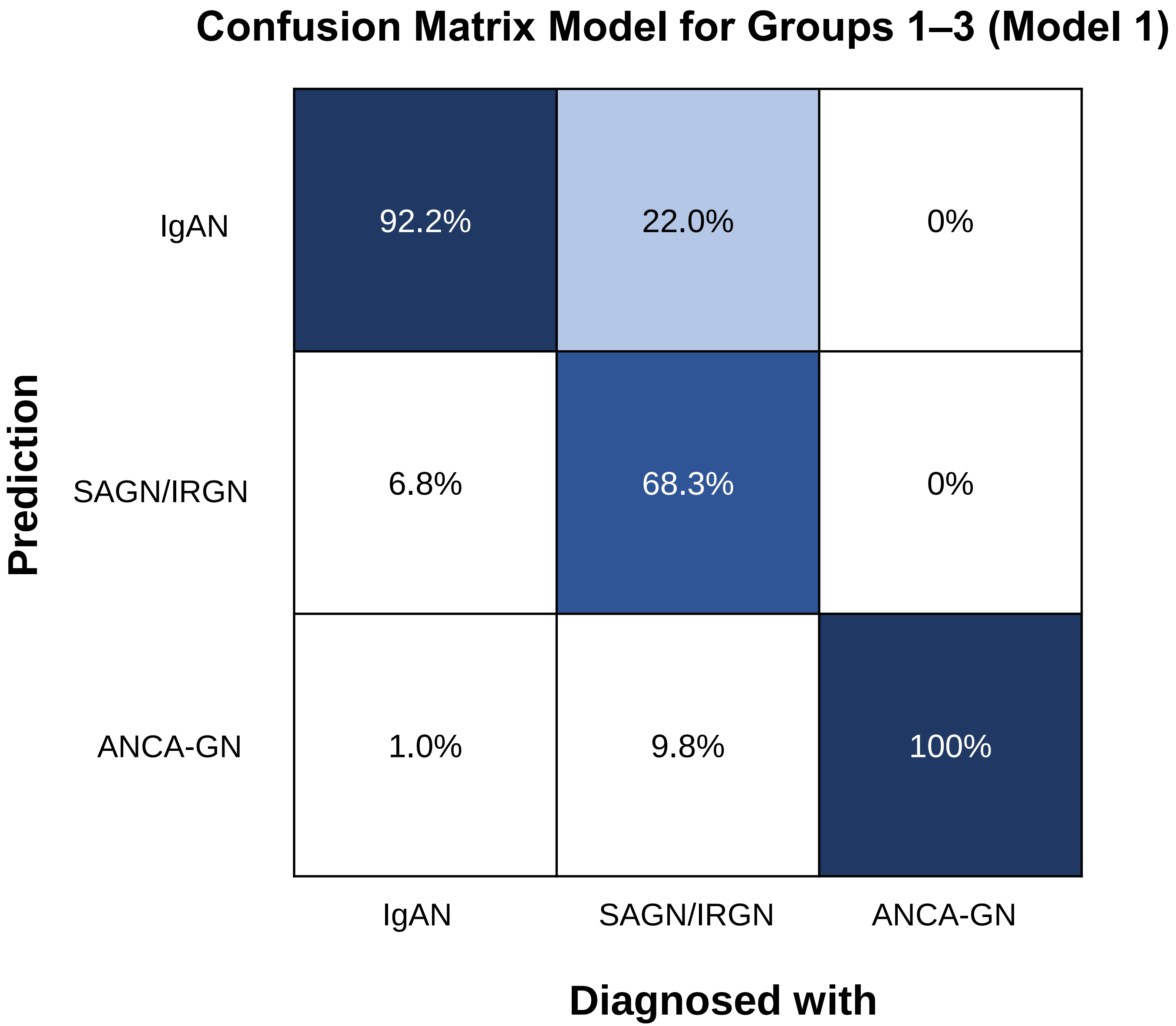


Supplementary Figure 1

B

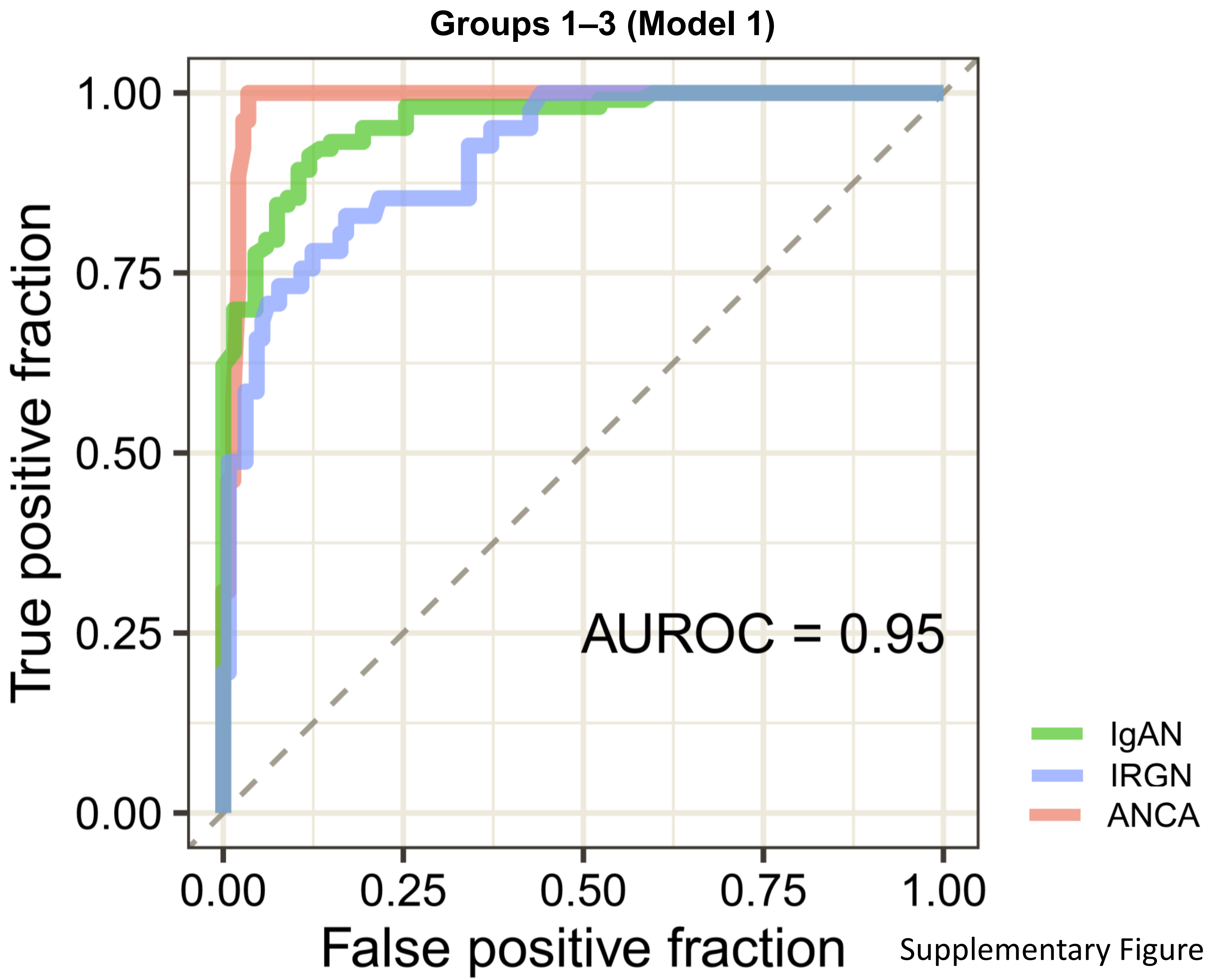


Supplementary Figure 1

C

### Confusion Matrix Model for Groups 1–3 (Model 3)

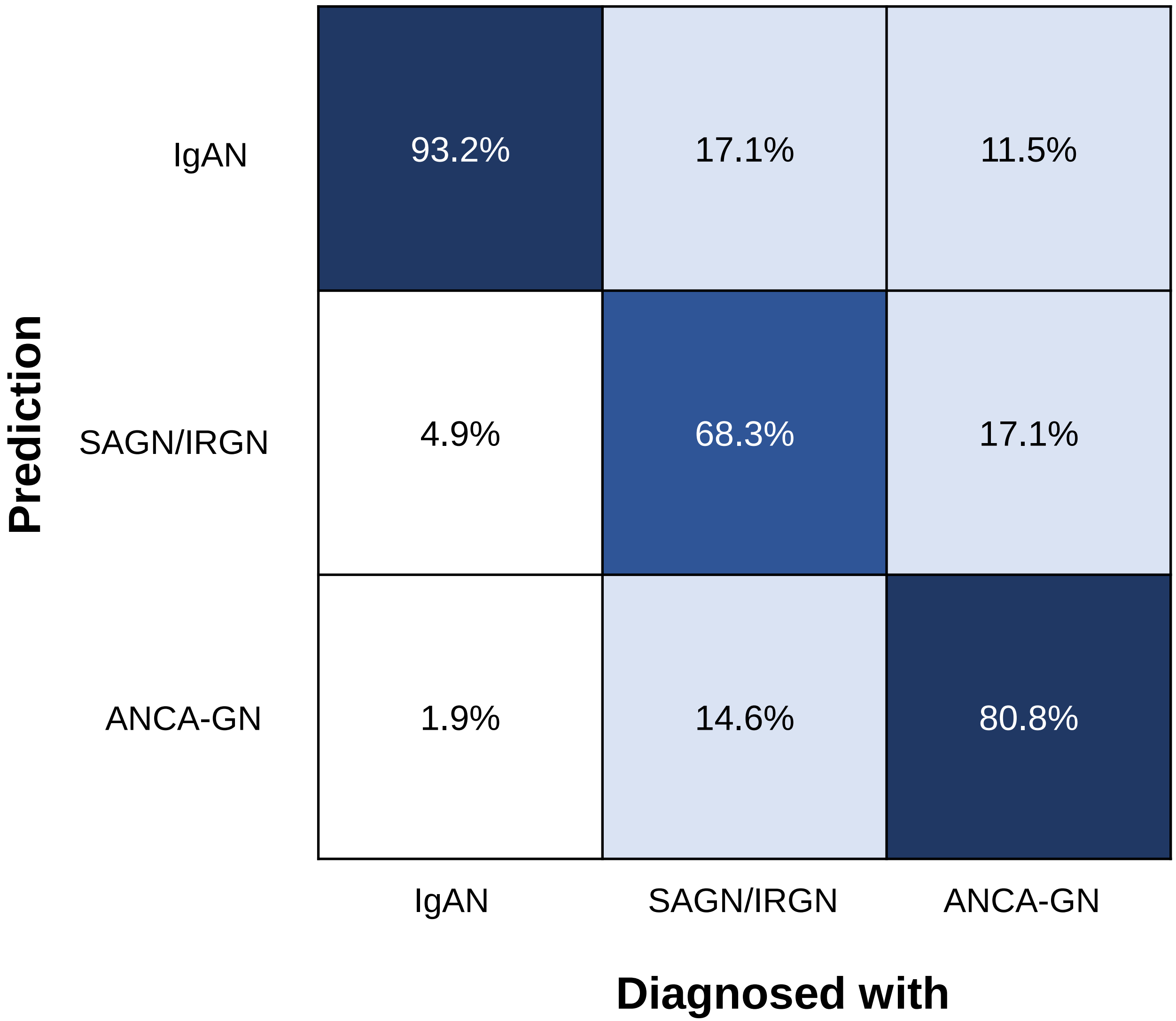


Supplementary Figure 1

D

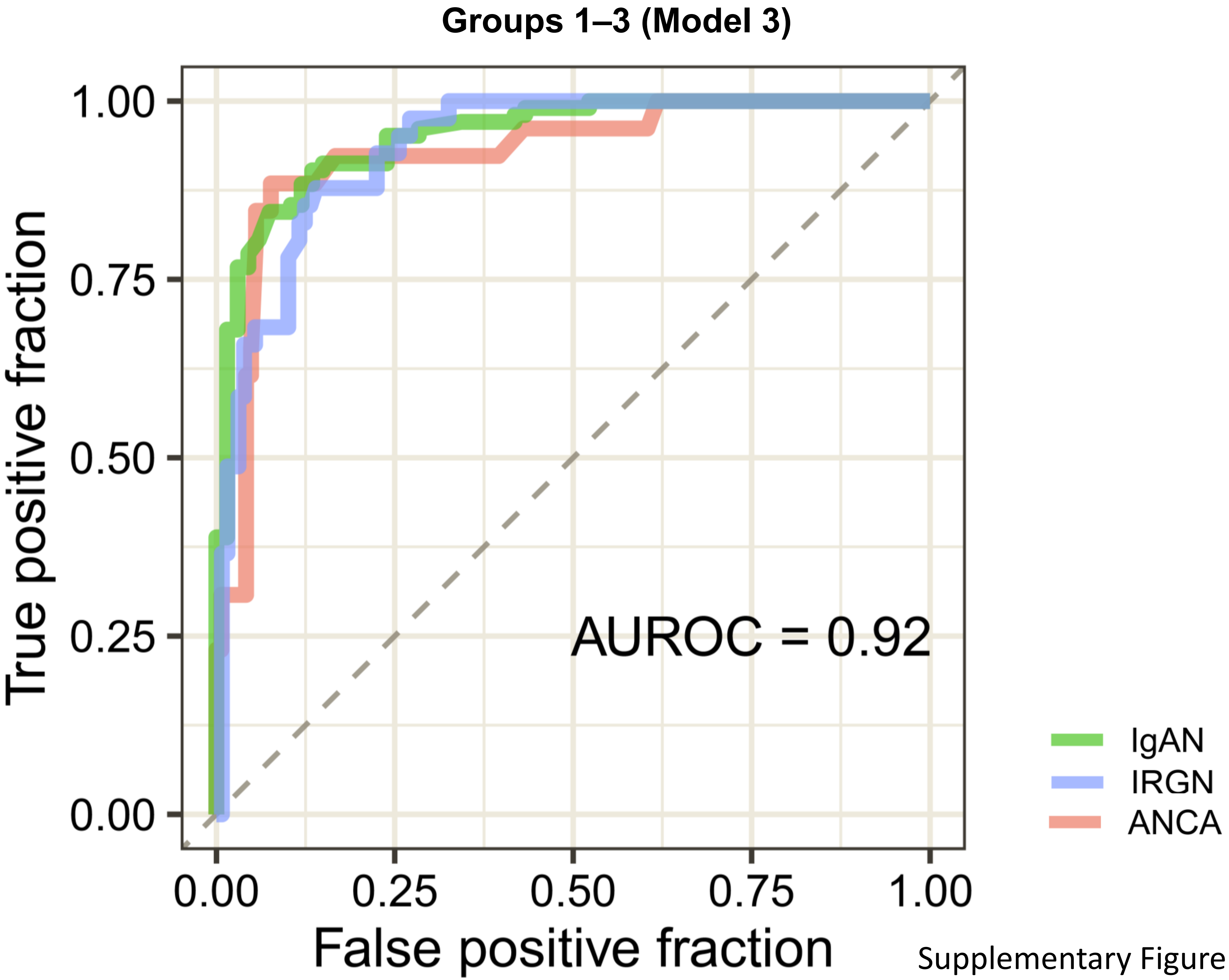


Supplementary Figure 1

A

### Confusion Matrix Model for Group X (Model 1)

| Prediction \ Treated as | “IgAN” | “SAGN/IRGN” | “ANCA-GN” |
|---|---|---|---|
| IgAN | 38.7% | 18.9% | 0% |
| SAGN/IRGN | 51.6% | 56.3% | 35.0% |
| ANCA-GN | 9.7% | 25.0% | 65.0% |

Supplementary Figure 2

B

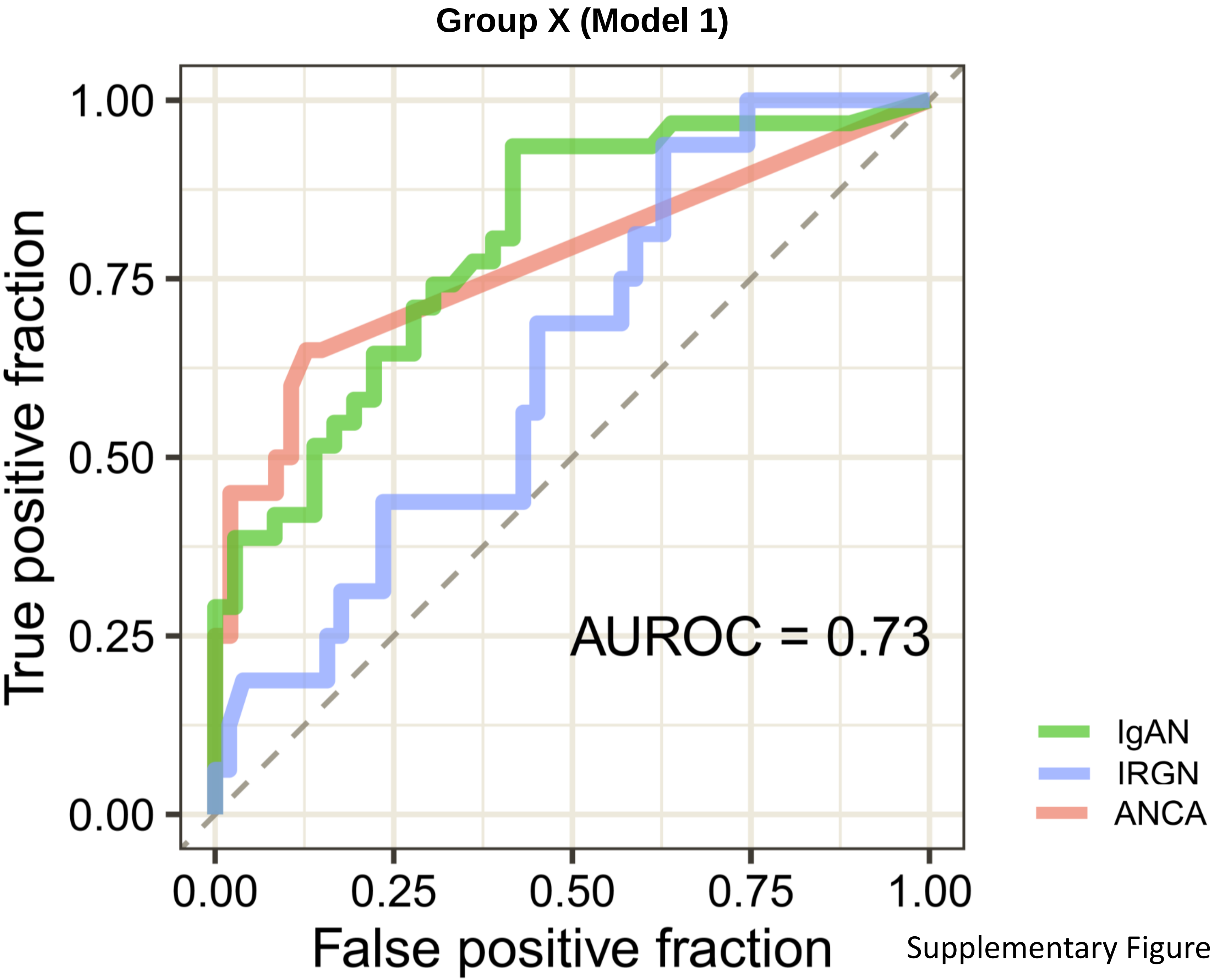


Supplementary Figure 2

C

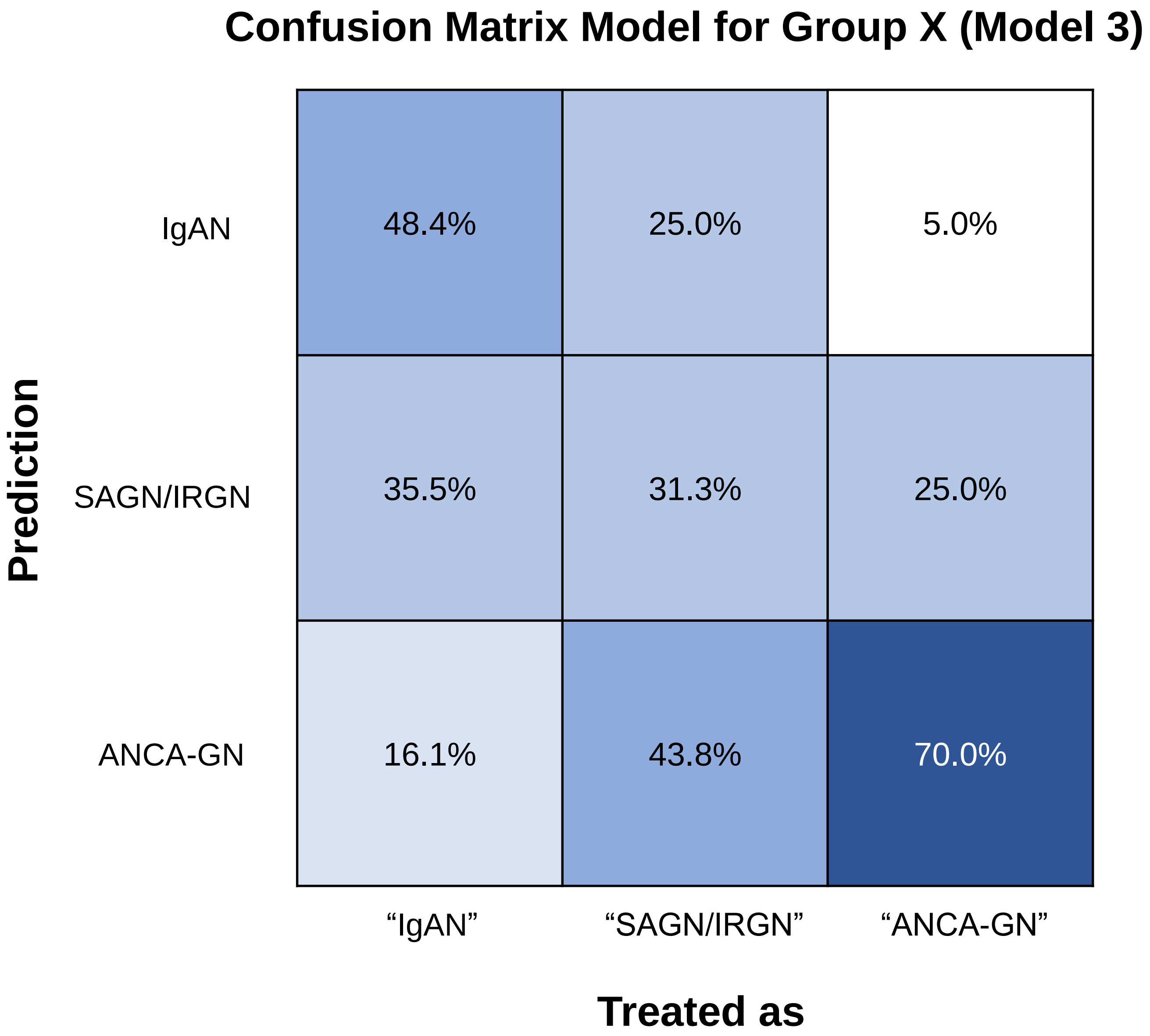


Supplementary Figure 2

D

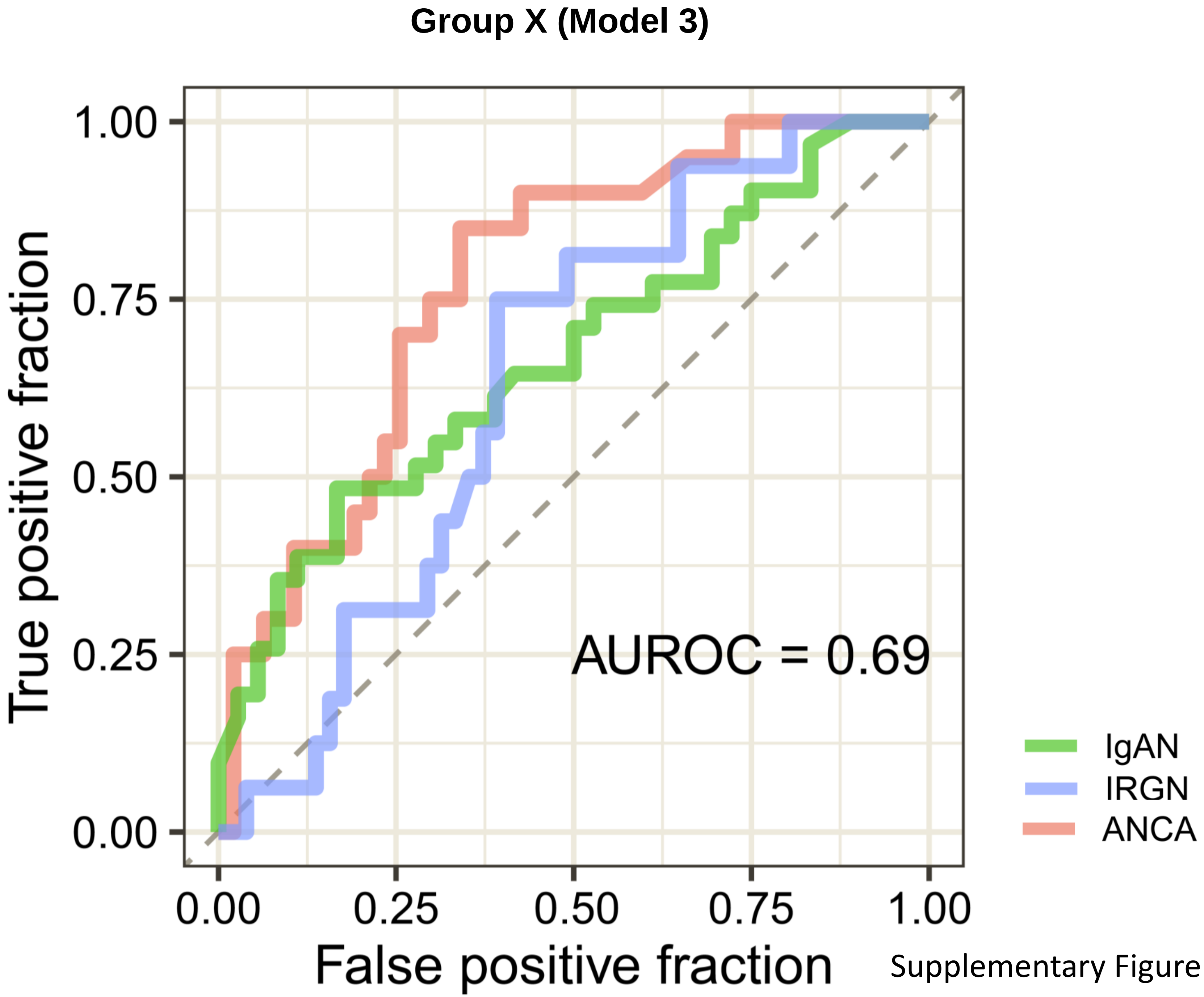


Supplementary Figure 2

A

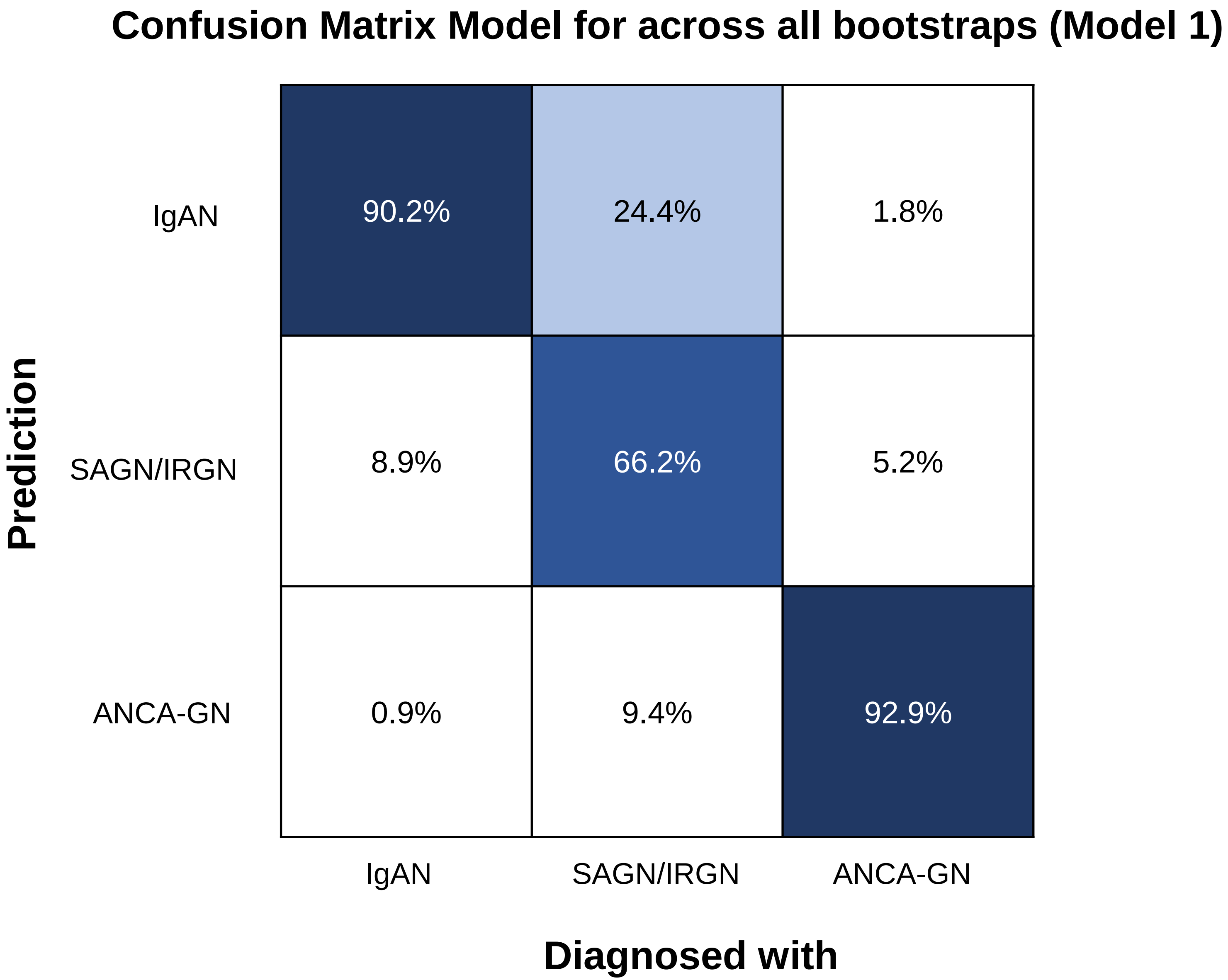


Supplementary Figure 3

B

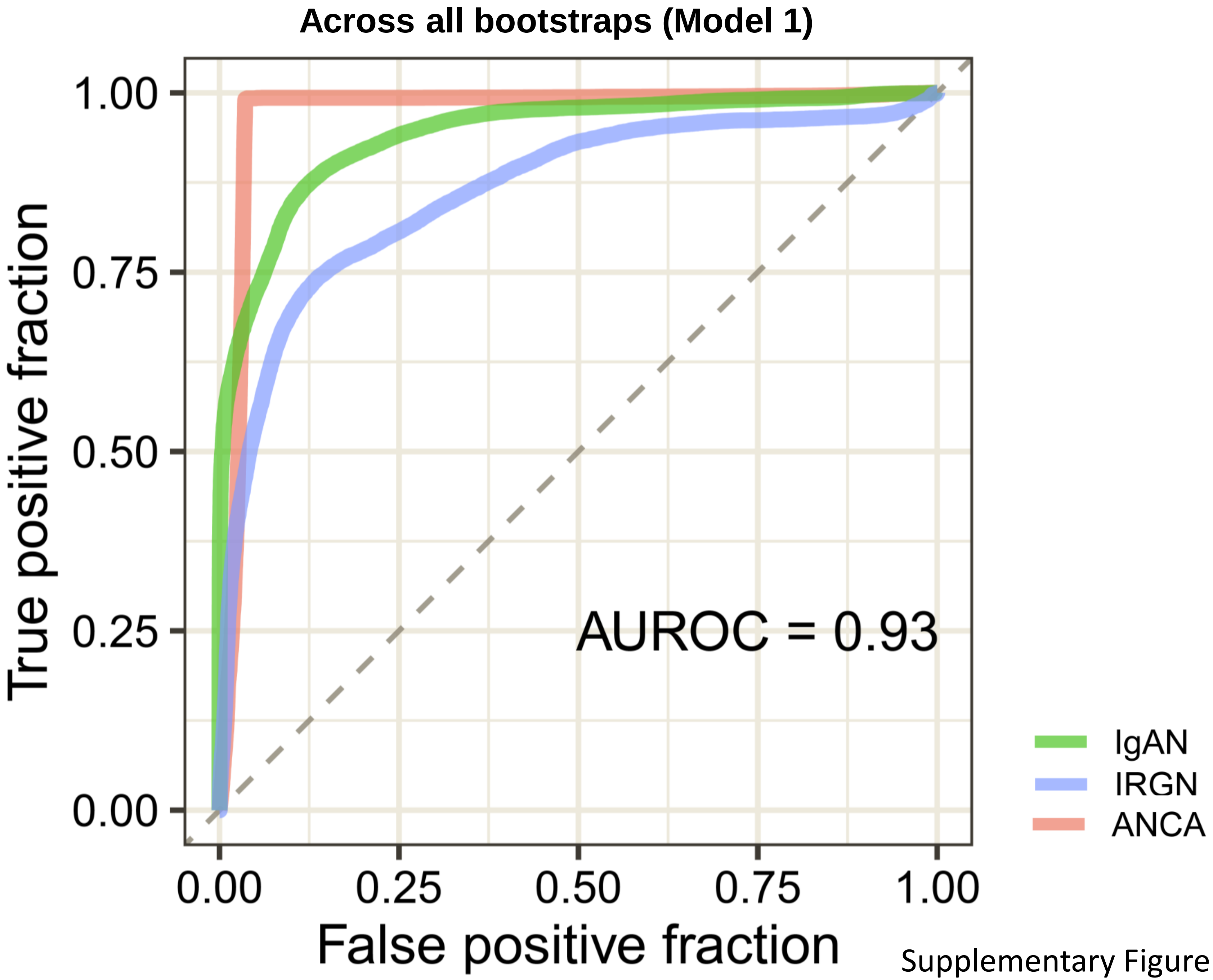


Supplementary Figure 3

C

**Confusion Matrix Model for across all bootstraps (Model 2)**

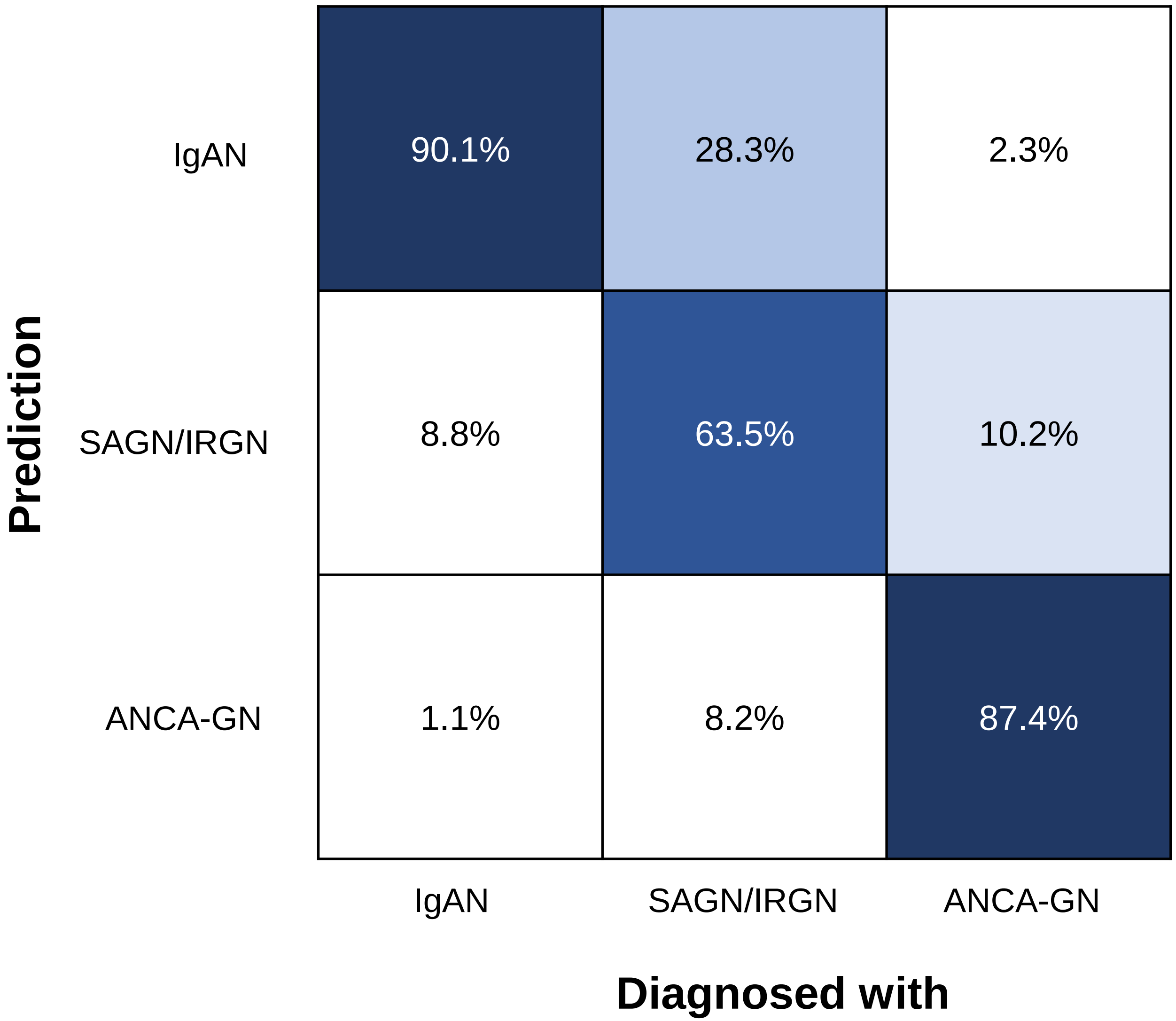


Supplementary Figure 3

D

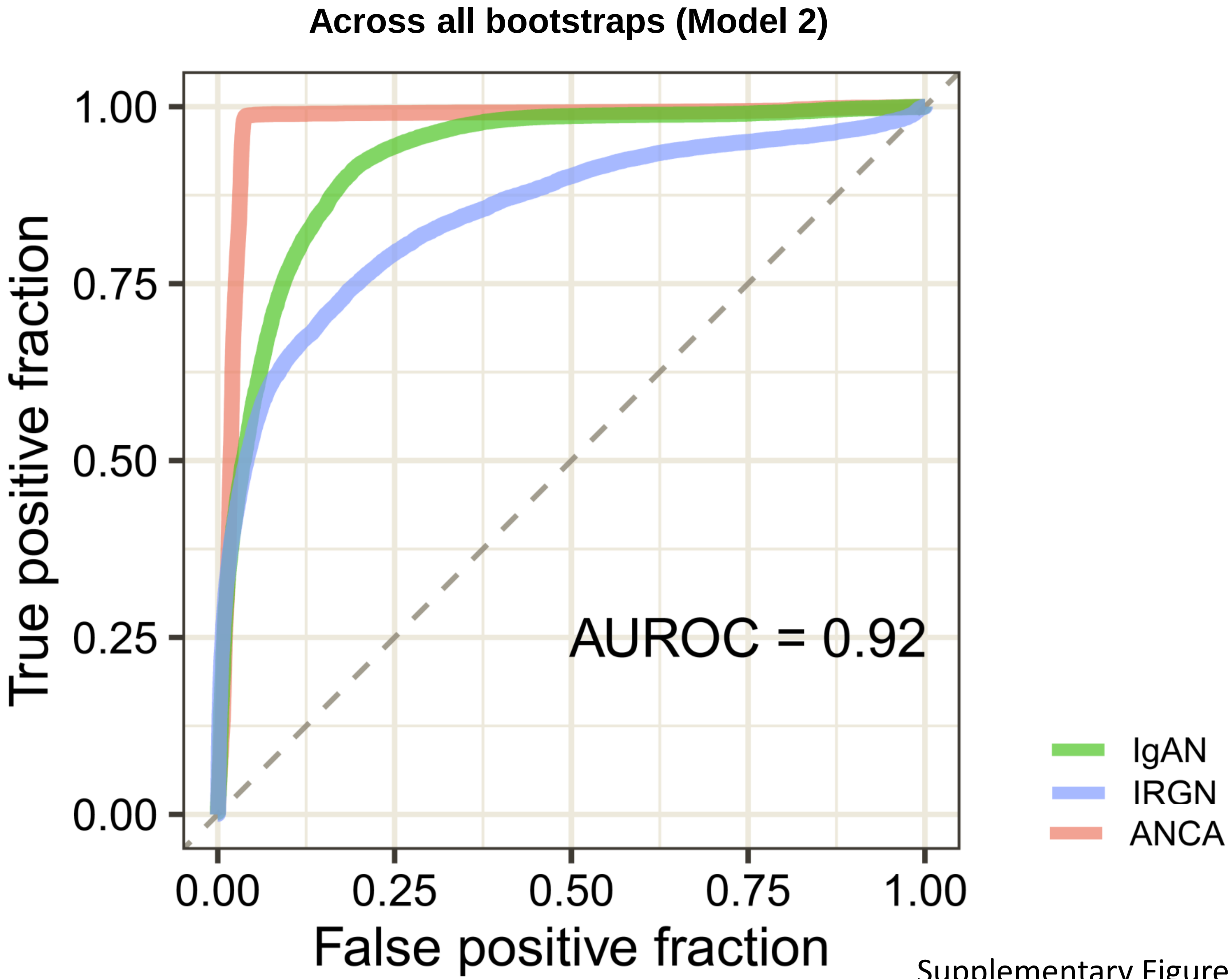


Supplementary Figure 3

E

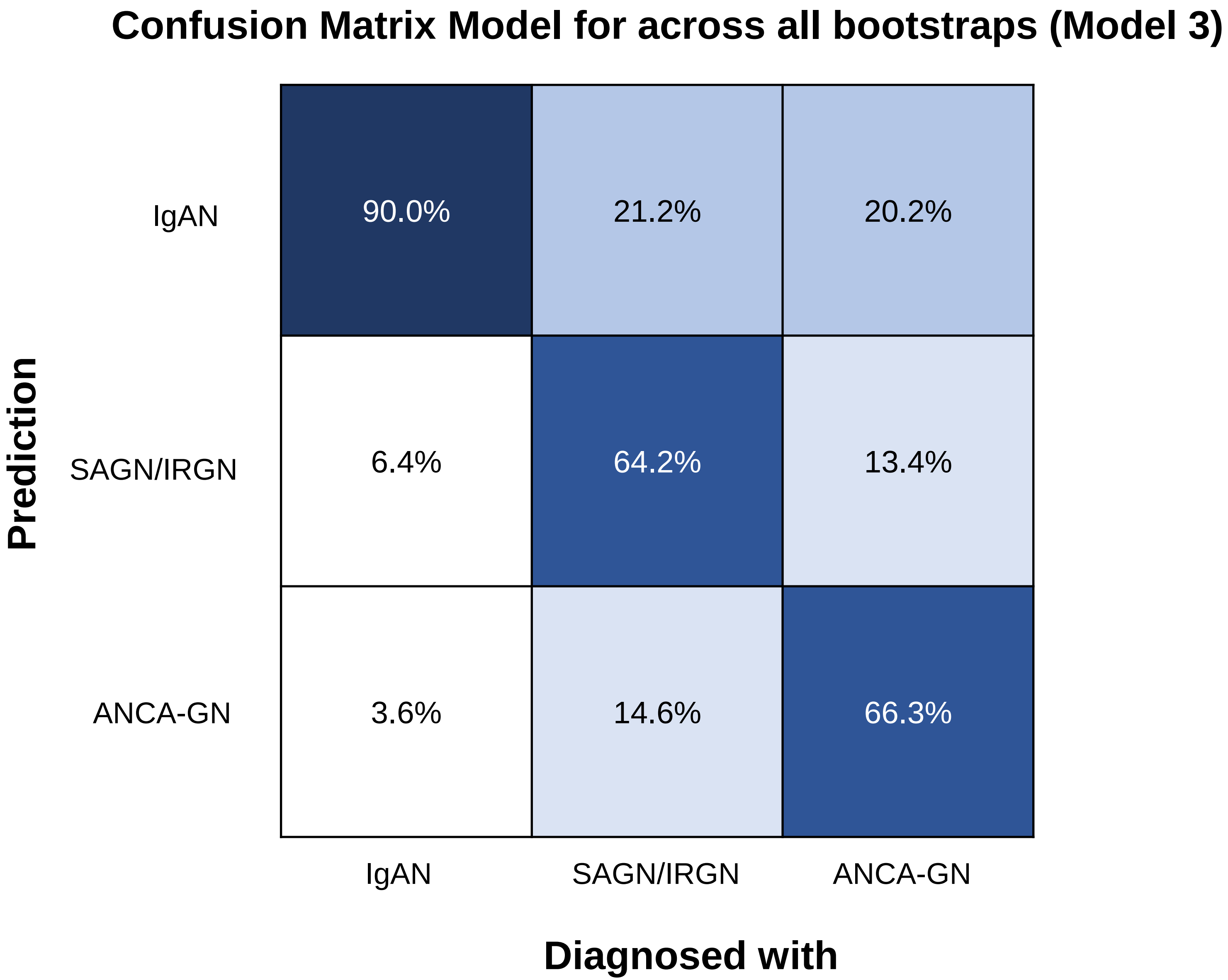


Supplementary Figure 3

F

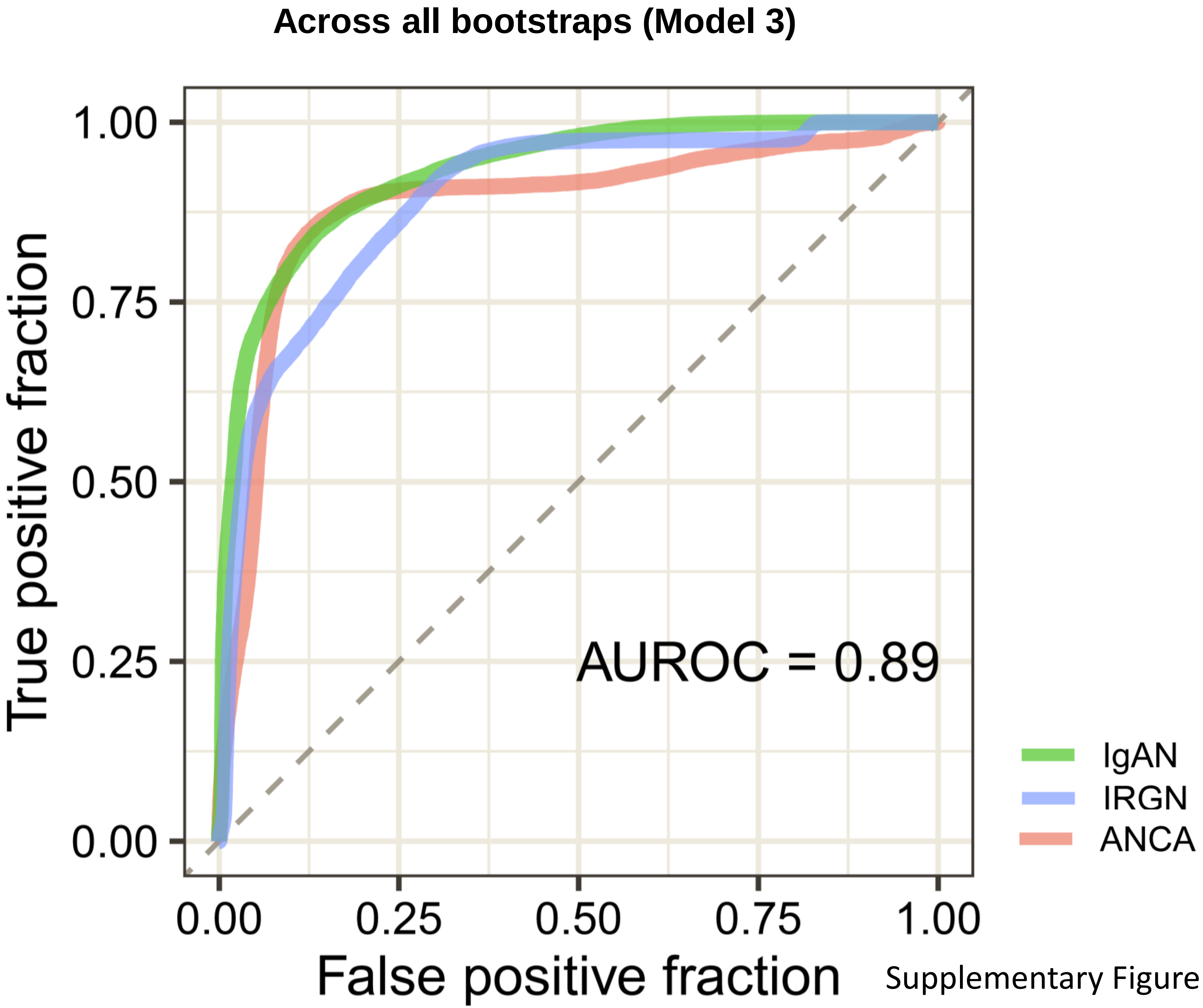


Supplementary Figure 3
