## Supplementary Tables for "Glomerular crescents, IgA-deposits, ANCA, infection – unravelling the diagnostic conundrum"

**Supplementary materials**

**Supplementary Table 1. Selected candidate parameters for prediction models**
**Supplementary Table 2. Oxford classification for Group 1**
**Supplementary Table 3. Details of infections in the SAGN/IRGN group (Group 2)**
**Supplementary Table 4. Coefficients of the variables for the three models**
**Supplementary Table 5. Summary of AUROC in the three models**
**Supplementary Table 6. Accuracy of the prediction models (Group 1-3)**
**Supplementary Table 7. Follow up summary of cases managed as "infection related glomerulonephritis" in Group X**
**Supplementary Table 8. Difference between Groups 1–3 and followed up cases in Group X at 1Y**

**Supplementary Figure 1. Confusion Matrix and ROC curves of Model 1, and 3 for Group 1-3**
**Supplementary Figure 2. Confusion Matrix and ROC curves of Models 1–3 for all bootstrap models.**
**Supplementary Figure 3. Confusion Matrix and ROC curves of Model 1, and 3 for Group X**

**Supplementary Table 1. Selected candidate parameters for prediction models**

| |
|---|
| **Clinical parameters** |
| antineutrophil cytoplasmic antibodies serology at the time of biopsy (Yes, No, Not available) |
| serum low C3 (Yes, No, Not available) |
| age (years old) |
| serum creatinine levels (mg/dL) at the time of biopsy (not available cases are excluded) |
| **Pathological parameters** |
| Cellular crescent % (continuous value) |
| Fibrous + fibrocellular crescent % (continuous value) |
| Segmental sclerosis (Yes, No) |
| Intracapillary hypercellularity (Yes, No) |
| IgA intensity in immunofluorescence (0-3+) |
| C3 intensity in immunofluorescence (0-3+) |

Candidate ten parameters were selected based on the clinicopathological significance in differentiating cases with IgA positive on glomeruli and crescents (Clin J Am Soc Nephrol 12: 39–49, 2017)

**Supplementary Table 2. Oxford classification for IgA nephropathy (Group 1)**

| | **M** | **E** | **S** | **T** | **C** |
|---|---|---|---|---|---|
| 0 (n,%) | 26, 24% | 60, 56% | 46, 43% | 41, 38% | 17, 16% |
| 1 (n,%) | 82, 76% | 48, 44% | 62, 57% | 45, 42% | 74, 69% |
| 2 (n,%) | | | | 22, 20% | 14, 16% |

Oxford classification was done described as previously (1)


1. Haas M, Verhave JC, Liu ZH, Alpers CE, Barratt J, Becker JU, Cattran D, Cook HT, Coppo R, Feehally J, Pani A, Perkowska-Ptasinska A, Roberts ISD, Soares MF, Trimarchi H, Wang S, Yuzawa Y, Zhang H, Troyanov S, Katafuchi R: A multicenter study of the predictive value of crescents in IgA nephropathy. *J Am Soc Nephrol* 28: 691–701, 2017

**Supplementary Table 3. Details of infections in the SAGN/IRGN group (Group 2)**

| **Infection site** | **Number of patients** |
|---|---|
| Infectious endocarditis | 15 |
| sepsis bacteremia etiology unknown | 6 |
| pneumonia | 6 |
| skin infections | 6 |
| cellulitis | 5 |
| osteomyelitis | 4 |
| abscess | 4 |
| infectious arthritis | 2 |
| thoracic spine infection | 1 |
| trauma | 1 |
| colitis | 1 |
| **Pathogenic organisms** | |
| **Genus of Staphylococcus** | |
| MRSA | 26 |
| MSSA | 6 |
| *Staphylococcus hominis* | 1 |
| *Staphylococcus epidermidis* | 1 |
| MRCNS | 1 |
| **Genus of Streptococcus** | |
| *Streptococcus viridans* | 2 |
| *Streptococcus mitis* | 1 |
| *Streptococcus group F* | 1 |
| *Group G streptococcus* | 1 |
| *Streptococcus species* | 1 |
| *Streptococcus mutans* | 1 |
| **Gram positive** | |
| *Enterococcus faecalis* | 2 |
| *Enterococcus group* | 1 |
| Gram positive cocci | 1 |
| **Others** | |
| *Pseudomonas aeruginosa* | 2 |
| *Klebsiella* | 2 |
| *Enterobacter* | 2 |
| *Clostridium perfringens* | 1 |
| *Candida* | 1 |

Duplicated cases are shown in this table. SAGN/IRGN, *Staphylococcus* infection associated glomerulonephritis / infection related glomerulonephritis, MRSA, methicillin-resistant staphylococcus aureus; MSSA, methicillin-sensitive staphylococcus aureus; MRCNS, methicillin resistant coagulase-negative staphylococci

**Supplementary Table 4. Coefficients of the variables for the three models**

**Model 1**

| Outcome level[1] | Term | Coefficient (95% confidence interval) | Standard error | p-value |
|---|---|---|---|---|
| IRGN | Intercept | 4.68 (1.41, 7.95) | 1.67 | 0.005 |
| IRGN | IgA intensity | -1.96 (-2.91, -1.00) | 0.49 | <0.001 |
| IRGN | Cellular crescent (%) | 0.11 (0.03, 0.18) | 0.04 | 0.004 |
| IRGN | Confirmed not ANCA[2] | -2.42 (-5.40, 0.56) | 1.52 | 0.11 |
| IRGN | Not run for ANCA[2] | -1.80 (-4.75, 1.15) | 1.51 | 0.23 |
| IRGN | Segmental sclerosis present | -3.58 (-5.93, -1.23) | 1.20 | 0.003 |
| ANCA-GN | Intercept | 7.01 (2.78, 11.25) | 2.16 | 0.001 |
| ANCA-GN | IgA intensity | -2.64 (-4.56, -0.72) | 0.98 | 0.007 |
| ANCA-GN | Cellular crescent (%) | 0.11 (0.03, 0.20) | 0.04 | 0.009 |
| ANCA-GN | Confirmed not ANCA | $1.76*10^{-7}$ (N/A, N/A) | N/A | N/A |
| ANCA-GN | Not run for ANCA | $3.51*10^{-8}$ (N/A, N/A) | N/A | N/A |
| ANCA-GN | Segmental sclerosis present | -2.18 (-5.40, 1.04) | 1.64 | 0.19 |

IRGN, infection related glomerulonephritis; IgAN, immunoglobulin A nephropathy; ANCA, antineutrophil cytoplasmic antibodies; N/A, not available

[1] The reference category is IgAN.

[2] ANCA is run at the clinician's discretion. The reference category is confirmed high ANCA. All patients with ANCA as an outcome were confirmed low ANCA.

**Model 2**

| Outcome level[1] | Term | Coefficient (95% confidence interval) | Standard error | p-value |
|---|---|---|---|---|
| IRGN | Intercept | 111.14 (3.09, 3992.36) | 1.83 | 0.01 |
| IRGN | IgA intensity | 0.12 (0.05, 0.31) | 0.47 | <0.001 |
| IRGN | Cellular crescent (%) | 1.12 (1.05, 1.20) | 0.03 | 0.001 |
| IRGN | Confirmed not ANCA[2] | 0.11 (0.004, 3.25) | 1.72 | 0.20 |
| IRGN | Not run for ANCA[2] | 0.19 (0.01, 5.35) | 1.71 | 0.33 |
| IRGN | FC+F crescent (%) | 0.90 (0.84, 0.97) | 0.04 | 0.005 |
| ANCA-GN | Intercept | 358.02 (3.75, 34,143) | 2.33 | 0.01 |
| ANCA-GN | IgA intensity | 0.05 (0.01, 0.48) | 1.15 | 0.01 |
| ANCA-GN | Cellular crescent (%) | 1.15 (1.05, 1.25) | 0.04 | 0.002 |
| ANCA-GN[2] | Confirmed not ANCA | $5.12*10^{-6}$ (N/A, N/A) | N/A | N/A |
| ANCA-GN | Not run for ANCA | 61.60 (N/A, N/A) | N/A | N/A |
| ANCA-GN | FC+F crescent (%) | 1.03 (0.93, 1.14) | 0.05 | 0.60 |

IRGN, infection related glomerulonephritis; IgA, immunoglobulin A; ANCA, antineutrophil cytoplasmic antibodies; N/A, not available; FC+F crescent, fibrocellular crescent+ fibrous crescent

[1] The reference category is IgA nephropathy.

[2] ANCA is run at the clinician's discretion. The reference category is confirmed high ANCA. All patients with ANCA as an outcome were confirmed low ANCA.

**Model 3:**

| Outcome level[1] | Term | Coefficient (95% confidence interval) | Standard error | p-value |
|---|---|---|---|---|
| IRGN | Intercept | 2.70 (0.56, 4.85) | 1.09 | 0.01 |
| IRGN | IgA intensity | -2.95 (-4.21, -1.69) | 0.64 | <0.001 |
| IRGN | Cellular crescent (%) | 0.11 (0.05, 0.18) | 0.03 | <0.001 |
| IRGN | Endocapillary hypercellularity present | 3.12 (1.65, 4.59) | 0.75 | <0.001 |
| IRGN | Segmental sclerosis present | -4.04 (-6.34, -1.75) | 1.17 | <0.001 |
| ANCA-GN | Intercept | 4.62 (2.37, 6.87) | 1.15 | <0.001 |
| ANCA-GN | IgA intensity | -4.11 (-5.62, -2.61) | 0.77 | <0.001 |
| ANCA-GN | Cellular crescent (%) | 0.12 (0.06, 0.19) | 0.03 | <0.001 |
| ANCA-GN | Endocapillary hypercellularity present | -0.82 (-2.75, 1.10) | 0.98 | 0.40 |
| ANCA-GN | Segmental sclerosis present | -0.14 (-1.59, 1.31) | 0.74 | 0.85 |

IRGN, infection related glomerulonephritis; IgA, immunoglobulin A; ANCA, antineutrophil cytoplasmic antibodies

[1] The reference category is IgA nephropathy.

**Supplementary Table 5. Summary of area under the receiver operating curve in the three models**

| Model and parameters | Groups 1–3 | Across all bootstraps | Group X |
|---|---|---|---|
| **[Model 1]**: cellular crescent%, IgA intensity, ANCA serology, and segmental sclerosis | 0.95 | 0.93 | 0.73 |
| **[Model 2]**: cellular crescent%, IgA intensity, ANCA serology, and fibrous and fibrocellular crescent% | 0.95 | 0.92 | 0.75 |
| **[Model 3]**: cellular crescent%, IgA intensity, fibrous and fibrocellular crescent%, and endocapillary hypercellularity | 0.92 | 0.89 | 0.69 |

IgA, immunoglobulin A; ANCA, antineutrophil cytoplasmic antibodies

**Supplementary Table 6. Accuracy of the prediction models (Group 1-3)**

| Model and parameters | IgAN (Group 1) (n=103) | SAGN/IRGN (Group 2) (n=41) | ANCA-GN (Group 3) (n=26) | AUROC |
|---|---|---|---|---|
| **[Model 1]**: cellular crescent%, IgA intensity, ANCA serology, and segmental sclerosis | 95, 92% | 28, 68% | 26, 100% | 0.95 |
| **[Model 2]**: cellular crescent%, IgA intensity, ANCA serology, and fibrous and fibrocellular crescent% | 96, 93% | 27, 66% | 26, 100% | 0.95 |
| **[Model 3]**: cellular crescent%, IgA intensity, fibrous and fibrocellular crescent%, and endocapillary hypercellularity | 96, 93% | 28, 68% | 21, 81% | 0.92 |

Five patients in Group 1 and two patients in Group 2 were excluded from constructing prediction models because their serum creatinine levels (one of the candidate parameters) were not available at the time of biopsy.

IgAN, immunoglobulin A nephropathy; SAGN/IRGN, *Staphylococcus* infection–associated glomerulonephritis/ infection related glomerulonephritis; ANCA-GN, antineutrophil cytoplasmic antibodies associated glomerulonephritis; AUROC, area under the receiver operating curve

**Supplementary Table 7. Follow up summary of cases managed as "infection related glomerulonephritis" in Group X**

| Pt No. | Age | Sex | sCr at bx (mg/dL) | infections | pathogens discovered after bx | treatment | comorbidities | renal function | alive or death at 1 year |
|---|---|---|---|---|---|---|---|---|---|
| 1 | 51-60 | M | 5.9 | cellulitis | not determined | antibiotics | DM, HT, HCV+, morbidly obese | recovered | alive |
| 2 | 71-80 | M | 6.6 | pneumonia | not determined | antibiotics and steroids | CAD, heat failure, COPD, morbidly obese | renal death | alive |
| 3 | 71-80 | M | 1.1 | pressure ulcers paraplegic | MRSA | antibiotics | HT, PAD, carotid artery stenosis, AAA, history of osteomyelitis. | unchanged | alive |
| 4 | 61-70 | F | 15.0 | pneumonia | not determined | antibiotics | HT, hyperlipidemia, leukocytosis | renal death | alive |
| 5 | 61-70 | M | 4.7 | vertebral infection | MRSA | antibiotics | HT, prior history of pneumonia | renal death | alive |
| 6 | 51-60 | M | 2.5 | gastrointestinal infections | not determined | supportive | DM, HT, morbidly obese, pancreatitis, liver cirrhosis | unchanged | death |
| 7 | 31-40 | M | 3.0 | cellulitis | MRSA | antibiotics | Crohn's disease alcoholism, heart failure, cardiomyopathy | renal death | death |
| 8 | 41-50 | F | 6.3 | pneumonia | not determined | antibiotics and steroid | DM, HT, HCV+, morbidly obese | unknown | unknown |
| 9 | 51-60 | M | 3.8 | skin infection | not determined | antibiotics and steroid | C-ANCA positive | recovered | death |
| 10 | 41-50 | M | 3.7 | foot infection | not determined | antibiotics | DM, HT, HCV+, morbidly obese | recovered | alive |
| 11 | 51-60 | M | 4.4 | skin infection | not determined | antibiotics | DM, HT, morbidly obese | recovered | alive |
| 12 | 71-80 | M | 2.5 | endocarditis | *Bartonella henselae* | antibiotics | Splenomegaly, hepatomegaly, RF+ | recovered | alive |
| 13 | 51- | M | 6.5 | cellulitis | not determined | antibiotics | Morbidly obese, HT, ANCA+ | renal death | alive |

| | | | | | | | | | |
|---|---|---|---|---|---|---|---|---|---|
| | 60 | | | | | | | | |
| 14 | 61-70 | M | 7.9 | cellulitis | not determined | antibiotics | DM, HT | unknown | death |
| 15 | 61-70 | M | 6.7 | cellulitis | not determined | supportive | CAD, unilateral kidney atrophy, BPH, HCV+ | renal death | alive |
| 16 | 61-70 | M | 3.5 | pneumonia | not determined | antibiotics and steroid | HT, ANCA+ | renal death | death |

sCr at bx, serum creatinine levels at the time of the biopsy; M, male; F, female; IRGN, infection related glomerulonephritis; MRSA, methicillin-resistant staphylococcus aureus; DM, diabetes mellites; HT, hypertension; HCV, hepatitis C virus; ANCA, antineutrophil cytoplasmic antibody; RF, rheumatoid factor; CAD, coronary artery disease; COPD, chronic obstructive pulmonary disease; PAD, Peripheral artery disease; AAA, abdominal aortic aneurysm; HCC, HCC, hepatocellular carcinoma; BPH, benign prostatic hyperplasia

**Supplementary Table 8. Difference between Groups 1–3 and followed up cases in Group X at 1Y**

| | IgAN (Group 1) (n=108) | "IgAN" (Group X) (n=31) | p value | SAGN/IRGN (Group 2) (n=43) | "SAGN/IRGN" (Group X) (n=16) | p value | ANCA-GN (Group 3) (n=26) | "ANCA-GN" (Group X) (n=20) | p value |
|---|---|---|---|---|---|---|---|---|---|
| **Age (year)** [a] | 40±16 | 59±16 | <0.001 | 53±15 | 60±11 | 0.09 | 59±18 | 60±14 | 0.95 |
| **Age range** | (7–86) | (30–84) | | (24–83) | (40–74) | | (24–87) | (29–91) | |
| **Male: Female** | 69:39 | 22:9 | 0.46 | 31:12 | 14:2 | 0.24 | 16:10 | 8:12 | 0.15 |
| **Caucasian** | 80, 83% (n=96) | 23, 85% (n=27) | 0.82 | 34, 94% (n=36) | 12, 92% (n=13) | 0.79 | 17, 81% (n=21) | 15, 88% (n=17) | 0.54 |
| **History of diabetes** | 10, 9% | 7, 23% | 0.06 | 13, 30% | 4, 25% | 0.69 | 6, 23% | 2, 10% | 0.23 |
| **History of infection (UTI and URTI excluded)** | 0, 0% | 5, 16% | <0.001 | 43, 100% | 10, 63% | <0.001 | 1, 5% | 4, 20% | 0.08 |
| **ANCA+** | 1, 2% (n=56) | 3, 14% (n=21) | 0.04 | 4, 15% (n=26) | 4, 36% (n=11) | 0.17 | 26, 100% (n=26) | 14, 74% (n=19) | 0.002 |
| **sCr (mg/dL)** | 3.1±3.1 | 4.0±2.1 | <0.001 | 4.6±2.1 | 5.1±3.2 | 0.75 | 5.6±4.1 | 5.2±2.3 | 0.81 |
| **Low C3 (yes)** | 3, 5% (n=57) | 2, 8% (n=26) | 0.67 | 14, 39% (n=36) | 4, 29% (n=14) | 0.49 | 0, 0% (n=17) | 1, 6% (n=18) | 0.24 |
| **Low C4 (yes)** | 1, 2% (n=57) | 1, 4% (n=26) | 0.58 | 3, 8% (n=36) | 2, 14% (n=14) | 0.54 | 0, 0% (n=17) | 0, 0% (n=18) | 1.00 |
| **Proteinuria (yes)** | 96, 94% (n=102) | 25, 100% (n=25) | 0.10 | 35, 100% (n=35) | 13, 100% (n=13) | 1.00 | 18, 100% (n=18) | 18, 100% (n=15) | 1.00 |
| **Nephrotic proteinuria or >3+** | 49, 48% (n=102) | 18, 72% (n=25) | 0.03 | 21, 60% (n=35) | 8, 62% (n=13) | 0.92 | 3, 17% (n=18) | 4, 27% (n=15) | 0.48 |
| **Hematuria (yes)** | 86, 91% (n=94) | 24, 96% (n=24) | 0.42 | 34, 94% (n=36) | 14, 100% (n=14) | 0.25 | 22, 96% (n=23) | 18, 100% (n=18) | 0.28 |
| **M- hypercellularity present** | 91, 84% | 20, 65% | 0.02 | 34, 79% | 11, 69% | 0.42 | 3, 12% | 5, 25% | 0.23 |
| **E-hypercellularity present** | 48, 44% | 17, 55% | 0.31 | 32, 74% | 5, 31% | 0.03 | 2, 8% | 2, 10% | 0.78 |
| **Segmental sclerosis present** | 62, 57% | 4, 13% | <0.001 | 1, 2% | 0, 0% | 0.42 | 8, 31% | 3, 15% | 0.21 |
| **Sclerotic glomeruli (%)**[a] | 18 (5–39) | 21 (11–43) | 0.28 | 6 (0–26) | 10 (3–18) | 0.66 | 27 (7–41) | 19 (5–29) | 0.29 |
| **C-crescents (%)**[a] | 2 (0–7) | 13 (4–26) | <0.001 | 13 (6–28) | 16 (4–38) | 0.86 | 7 (0–38) | 23 (4–48) | 0.12 |
| **FC+F-crescents (%)**[a] | 6 (0–24) | 4 (0–19) | 0.30 | 0 (0–0) | 0 (0–0) | 0.75 | 19 (6–46) | 0 (0–17) | 0.003 |
| **Total crescents (%)**[a] | 11 (6–25) | 27 (8–33) | 0.02 | 13 (6–29) | 18 (6–38) | 0.77 | 48 (24–69) | 36 (21–53) | 0.22 |
| **≥5 neutrophils per glomerulus present** | 1, 1% | 5, 16% | 0.001 | 11, 26% | 0, 0% | 0.005 | 1, 4% | 1, 5% | 0.85 |
| **Necrotizing lesions present** | 12, 11% | 16, 52% | <0.001 | 21, 49% | 9, 56% | 0.61 | 15, 58% | 15, 75% | 0.22 |

| | IgAN (Group 1) (n=108) | "IgAN" (Group X) (n=31) | p value | SAGN/IRGN (Group 2) (n=43) | "SAGN/IRGN" (Group X) (n=16) | p value | ANCA-GN (Group 3) (n=26) | "ANCA-GN" (Group X) (n=20) | p value |
|---|---|---|---|---|---|---|---|---|---|
| **IF/TA grade (0–3)[a]** | 1.7±1.0 | 1.6±0.8 | 0.39 | 1.1±0.9 | 1.2±0.5 | 0.47 | 1.7±0.9 | 1.7±0.6 | 0.51 |
| **ATN present** | 41, 38% | 19, 62% | 0.02 | 38, 88% | 15, 94% | 0.53 | 18, 69% | 16, 80% | 0.41 |
| **RBC cast present** | 31, 29% | 14, 45% | 0.09 | 29, 67% | 12, 75% | 0.57 | 13, 50% | 15, 75% | 0.08 |
| **IgG (0–3)[a]** | 0.5±0.6 | 0.7±0.7 | 0.14 | 0.9±0.8 | 0.6±0.6 | 0.19 | 0.6±0.6 | 1.0±0.9 | 0.13 |
| **IgA (0–3)[a]** | 2.4±0.6 | 1.8±0.7 | <0.001 | 1.5±0.6 | 1.5±0.5 | 0.76 | 1.1±0.6 | 1.3±0.6 | 0.23 |
| **C1q (0–3)[a]** | 0.3±0.4 | 0.2±0.4 | 0.58 | 0.4±0.6 | 0.2±0.3 | 0.14 | 0.2±0.3 | 0.1±0.3 | 0.55 |
| **C3 (0–3)[a]** | 1.6±0.8 | 1.5±0.7 | 0.47 | 1.9±0.7 | 1.9±0.8 | 0.88 | 1.2±0.7 | 1.2±0.5 | 0.99 |
| **κ (0–3)[a]** | 1.5±0.7 | 1.4±0.8 | 0.35 | 1.0±0.8 | 1.1±0.7 | 0.61 | 0.7±0.6 | 1.1±0.8 | 0.04 |
| **λ (0–3)[a]** | 2.1±0.7 | 1.6±0.7 | 0.001 | 1.2±0.7 | 1.2±0.7 | 0.94 | 0.8±0.5 | 1.2±0.7 | 0.11 |
| **C3 dominance present** | 10, 9% | 7, 22% | 0.06 | 20, 47% | 8, 50% | 0.81 | 12, 46% | 4, 20% | 0.17 |
| **λ dominance present** | 60, 57% | 13, 42% | 0.15 | 18, 42% | 4, 25% | 0.22 | 10, 38% | 6, 30% | 0.55 |
| **Only mesangial EDD present** | 58, 56% (n=104) | 9, 31% (n=29) | 0.02 | 8, 19% (n=43) | 4, 27% (n=15) | 0.52 | 11, 46% (n=24) | 9, 45% (n=20) | 0.96 |
| **Mesangial and capillary wall EDD present** | 45, 42% (n=104) | 18, 62% (n=29) | 0.07 | 34, 79% (n=43) | 11, 73% (n=15) | 0.65 | 4, 17% (n=24) | 7, 35% (n=20) | 0.16 |
| **Subepithelial humps present** | 6, 6% (n=104) | 4, 14% (n=29) | 0.18 | 9, 21% (n=43) | 3, 20% (n=15) | 0.94 | 0, 0% (n=24) | 1, 5% (n=20) | 0.21 |
| **Absence of EDD** | 1, 1% (n=104) | 2, 7% (n=29) | 0.09 | 1, 2% (n=43) | 0, 0% (n=15) | 0.44 | 9, 38% (n=24) | 4, 20% (n=20) | 0.20 |

[a] Numerical variable. Continuous values are shown with mean±standard deviation or median (interquartile range). Continuous values were analyzed using Wilcoxon sum test. Categorial values were analyzed using Chi-square test

IgAN, immunoglobulin A nephropathy; SAGN/IRGN, *Staphylococcus* infection–associated glomerulonephritis/ infection related glomerulonephritis; ANCAGN, antineutrophil cytoplasmic antibodies associated glomerulonephritis; M-hypercellularity, mesangial-hypercellularity; E-hypercellularity, endothelial hypercellularity; C-Crescent, cellular crescent; FC+F-Crescent, fibrocellular+fibrous crescent; IF/TA, interstitial fibrosis and tubular atrophy; ATN, acute tubular necrosis; RBC, red blood cell; EDD, electron dense deposit; UTI, urinary tract infections; URTI, upper respiratory tract infections
